# Pharmacokinetics of Cannabidiol: A systematic review and meta-regression analysis

**DOI:** 10.1101/2023.02.01.23285341

**Authors:** Ehsan Moazen-Zadeh, Alexandra Chisholm, Keren Bachi, Yasmin L. Hurd

## Abstract

**Background:** In this review, we provide an updated assessment of available evidence on the pharmacokinetics (PK) of cannabidiol (CBD) and explore the impact of different factors on PK outcomes.

**Materials and Methods:** This systematic review and meta-regression analysis was pre-registered (PROSPERO: CRD42021269857). We systematically searched Medline, Embase, PsychInfo, and Web of Science Core Collection up to November 19, 2022. Trials of CBD in healthy adults were included if they reported at least one of the PK parameters of interest, including Tmax, Cmax, AUC0-t, AUC0-inf, and T_1/2_, in serum or plasma. Studies of patient populations or CBD co-administration with other medications were excluded. The *National Heart, Lung, and Blood Institute’s Quality Assessment Tool for Before-After Studies with no Control Group* was used. Random-effects multivariable meta-regression analysis was conducted.

**Results:** A total of 112 trial arms from 39 studies were included; 26 trial arms had a “Good” quality, 70 “Fair,” and 16 “Poor.” Eight arms used inhalation CBD, 29 oromucosal, 73 oral, and 2 intravenous. CBD formulations could be categorized to nanotech (n=14), oil-based (n=21), alcohol-based (n=10), water-based (n=12), Sativex (n=17), and Epidiolex (n=22). For single-dose studies, CBD doses ranged between 2-100mg in inhalation, 5-50mg in oromucosal, and 0.42-6000mg in oral administration. Sixty-six trial arms had only male participants or a higher number of males than females. The duration of the PK session was between 4h-164h. A higher CBD dose was associated with higher Cmax, AUC0-t, and AUC0-inf. Compared to oral administration, oromucosal administration was associated with lower Cmax, AUC0-t, and AUC0-inf. Fed status was associated with higher Cmax and AUC0-t when compared to the fasting status. A higher ratio of female participants was associated with lower Tmax in oral administration and higher Cmax.

**Conclusion:** As expected, CBD dose, route of administration, and diet were major determinants of CBD pharmacokinetics with oral routes providing higher bioavailability and nanotechnology formulations a faster onset. Though CBD appeared to have a faster onset and longer duration in females, more studies are required to delineate the role of biological sex. Factors that influence CBD PK have implications for medication development and appropriate dosing in clinical practice.

## Introduction

Cannabidiol (CBD), a cannabinoid constituent of the cannabis plant, has exponentially gained attention in both research and clinical applications as a potential treatment of several neuropsychiatric and general medical conditions ^1^. Epidiolex® was the first FDA-approved plant-derived CBD medication. Today, many CBD-based formulations are in development aiming for FDA approval and numerous non-approved CBD preparations are available over the counter often implying beneficial ‘medicinal’ properties ^2^. However, many questions raised by clinicians, researchers and consumers of CBD products often relates to dosing and administration.

Pharmacokinetics (PK) is fundamental to medication development and guides appropriate dosing to achieve clinical effectiveness. Important factors normally considered for medication development include the route of administration and bioavailability. Keeping in line with the route of administration most preferred for medicinal purposes, the majority of CBD products currently available are for oral use. However, similar to other cannabinoids, CBD generally has poor bioavailability when consumed orally ^3^. That challenge has sparked a growth in the industry for the development of new nanotechnologies to improve bioavailability, thus increasing the diversity of formulations and delivery systems being used. There are now multiple CBD products being investigated in clinical studies with varying routes of administration, formulations and administration conditions. Data generated from such studies should help to shed significant light on CBD PK parameters relevant to identifying those products potentially most suitable for subsequent trials regarding CBD’s pharmacodynamic properties to alleviate specific clinical conditions. However, one of the major challenges across studies and even in previous reviews has been the incomparability of outcomes across different CBD PK studies due to the different units and scales of reporting PK parameters, mainly geometric versus arithmetic scales ^3^. While both scales have been commonly used in reporting PK data, they are not readily or precisely convertible which contributes to confusion regarding PK outcomes. Of the scales, geometric method is preferred for reporting certain PK parameters due to its greater robustness ^4^. No study has yet integrated the various CBD PK data on one scale.

In this review, we aimed to provide an updated systematic assessment of available evidence on the PK of CBD, covering recently published studies that were not included in previous reviews; provide comparable values of PK parameters from different studies on the same scale, and demonstrate patterns in outcomes based on the CBD dose and route of administration; and explore the simultaneous impact of different factors on PK outcomes using meta-regression models.

## Materials and Methods

### Protocol

This systematic review and meta-regression analysis was pre-registered on PROSPERO (https://www.crd.york.ac.uk/prospero/, ID: CRD42021269857) and conducted in accordance with PRISMA guidelines as much as it applied to pharmacokinetic studies.

### Research objectives and outcomes

This review systematically assessed PK studies of CBD (pure CBD or in combination with THC) in healthy adults for both the quality of the studies and PK outcomes. The primary outcome was patterns of PK parameters of interest classified based on the route of entry and CBD dose to find potential meaningful patterns in outcomes that could help predict the PK of CBD relevant for clinical applications. The secondary outcome was the statistical significance level for each of the variables/factors that could potentially influence PK parameters, based on the available literature resulting from meta-regression models.

There were five PK parameters of interest:

1. Tmax: time from CBD administration to the maximum concentration of CBD in plasma/serum
2. Cmax: maximum concentration of CBD in plasma/serum
3. AUC0-t: the area under the curve of serum/plasma concentrations plotted against time from CBD administration to a specific time-point, usually the end of the PK session
4. AUC0-inf: the extrapolation of AUC0-t to the infinity time point
5. Half-life, t_1/2_: the time needed for the concentration of the drug in the plasma to be reduced by 50%.

Potential variables of interest were: route of administration, CBD dose, CBD formulation, dietary (fasting/fed) status before CBD administration, abstinence from cannabis before CBD administration, sex, and duration of the PK session ^3,5,6^.

### Inclusion/exclusion criteria

Trials of any design were considered for inclusion if conducted on healthy adults (age between 18 to 65 years) and reported at least one of the PK parameters of interest after CBD administration in serum or plasma. CBD could be administered in any form, through any entry route, in any formulation or product. Studies were excluded if there was a history of major psychiatric disorders or general medical conditions in the sample, or if CBD was co-administered with another medication.

### Search strategy

We systematically searched Medline, Embase, PsychInfo, Web of Science Core Collection, LILACS, and OpenGrey from inception to September 19, 2021, and in the case of Google Scholar, for the first 200 citations, using the following search terms: [Title/abstract→ (CBD OR cannabidiol OR Sativex OR Epidiolex)] AND [Title/abstract→ (pharmacokinetic OR concentration OR serum OR plasma OR blood)]. The above search was systematically updated on November 19, 2022 to cover publications since 2021 to date. Search terms were in English, but no language or publication period restriction was applied. Appropriate special characters/suffixes were used to search for any extension of the above terms. After the systematic search and screening, previous reviews of PK studies on CBD were searched manually for relevant original studies in addition to the reference lists of original articles published in the past 5 years. Experts in the field were consulted to seek any missed literature that could be included.

### Study selection and data extraction

Two doctorate-level authors were co-trained and calibrated for the screening and data extraction on a sample of CBD PK studies. Any discrepancy would be discussed and resolved between the two screeners and the senior author at each stage. Conference abstracts and thesis reports were also included if the minimum required data was reported. Title-abstract and full-text screening were carried out in parallel using EndNote v. 9 (Cleverbridge, Inc., USA). Data extraction was carried out by one of the two authors, and then all the data was checked against the original source for accuracy by the other author. Microsoft Excel sheets with predefined columns were used for extracting data, including but not limited to full citation details, study design, participants’ age/sex/health status, sample size and dropouts, cannabis use pattern and abstinence status, fast/fed status before CBD administration, CBD source/supplier, details of CBD formulation, route of administration, CBD dosing, duration of the study session, values of any reported PK parameter in their original format, any relevant considerations of statistical methods, and finally any other relevant methodological information. Published papers, supplementary material, and pharmaceutical providers’ websites were all used to collect relevant information, and in some cases, authors were contacted to obtain further details.

### CBD formulation determination

CBD formulations were determined using three primary factors. First, when a patent was held and the specific ingredients were not provided in the methodology section for each study, a CBD sample would be designated as its own formulation (i.e., Epidiolex or Sativex). Second, when a methodology section indicated that the solution used a specific technology type (e.g., nanoparticle), these solutions were determined to be their own formulation. Third, when a study provided specific ingredients in the methodology section, the main base of the solution was determined to categorize different CBD formulation types. For example, most formulations consist mainly of oil-based, ethanol-based, gelatin-based, or water-based solutions. All CBD solutions were categorized using these three factors.

### Quality assessment

The two authors who carried out the screening and data extraction also conducted a parallel quality assessment of all the studies. Any differences in ratings would be discussed and resolved between the two screeners and the senior author. Given the lack of a widely-accepted quality assessment tool for PK studies, the US National Heart, Lung, and Blood Institute Quality Assessment Tool for Before-After Studies with no Control Group was deemed the best choice ^7^. The tool consists of 12 items assessing different aspects of before-after studies, from the clarity of the study question to the very end analysis. Each item is rated as Yes, No, Cannot Determine (CD), or Not Reported (NR). Specifically for the sample size question (Q5), a sample of >19 was considered as “Yes,” <10 was considered “No,” and in between was rated as “CD” ^8^. The assessor assigned an overall rating to each study as Poor, Fair, or Good. Toward a more precise and replicable application of this tool for PK studies, the authors weighted some items and developed a stratification strategy for overall rating, which can be found in supplementary material Section A.

### Narrative synthesis

Study characteristics were organized, summarized, and presented separately for each trial arm in Table 1 in the order of date of publication. Similarities and differences among trial arms were described, including characteristics of the population such as sex, cannabis use, study design, diet status, CBD formulations, CBD dose, and reported outcomes. Numerical values of PK parameters were presented in Supplement Table 1 and described in the quantitative synthesis section.

**Table 1.**
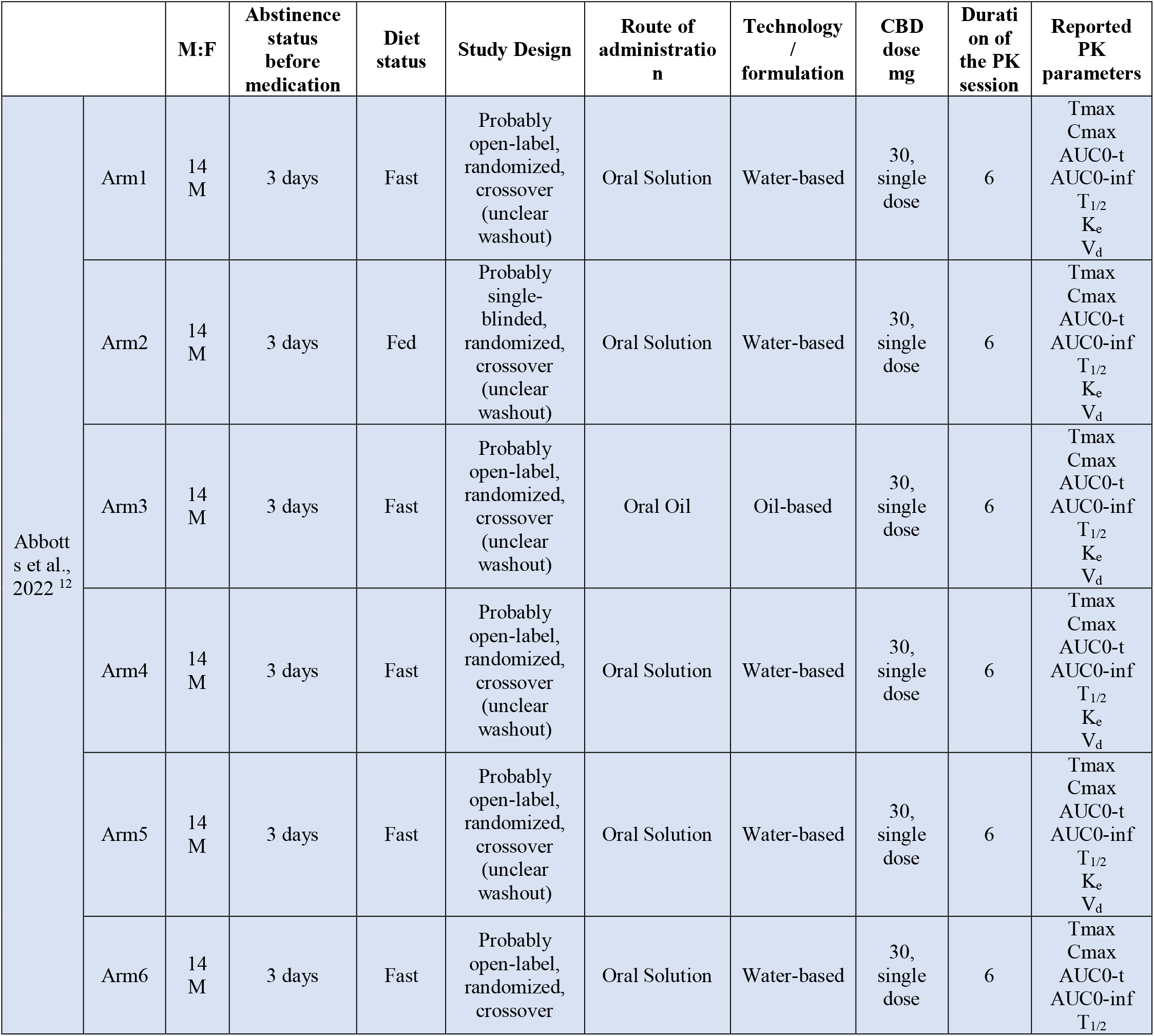
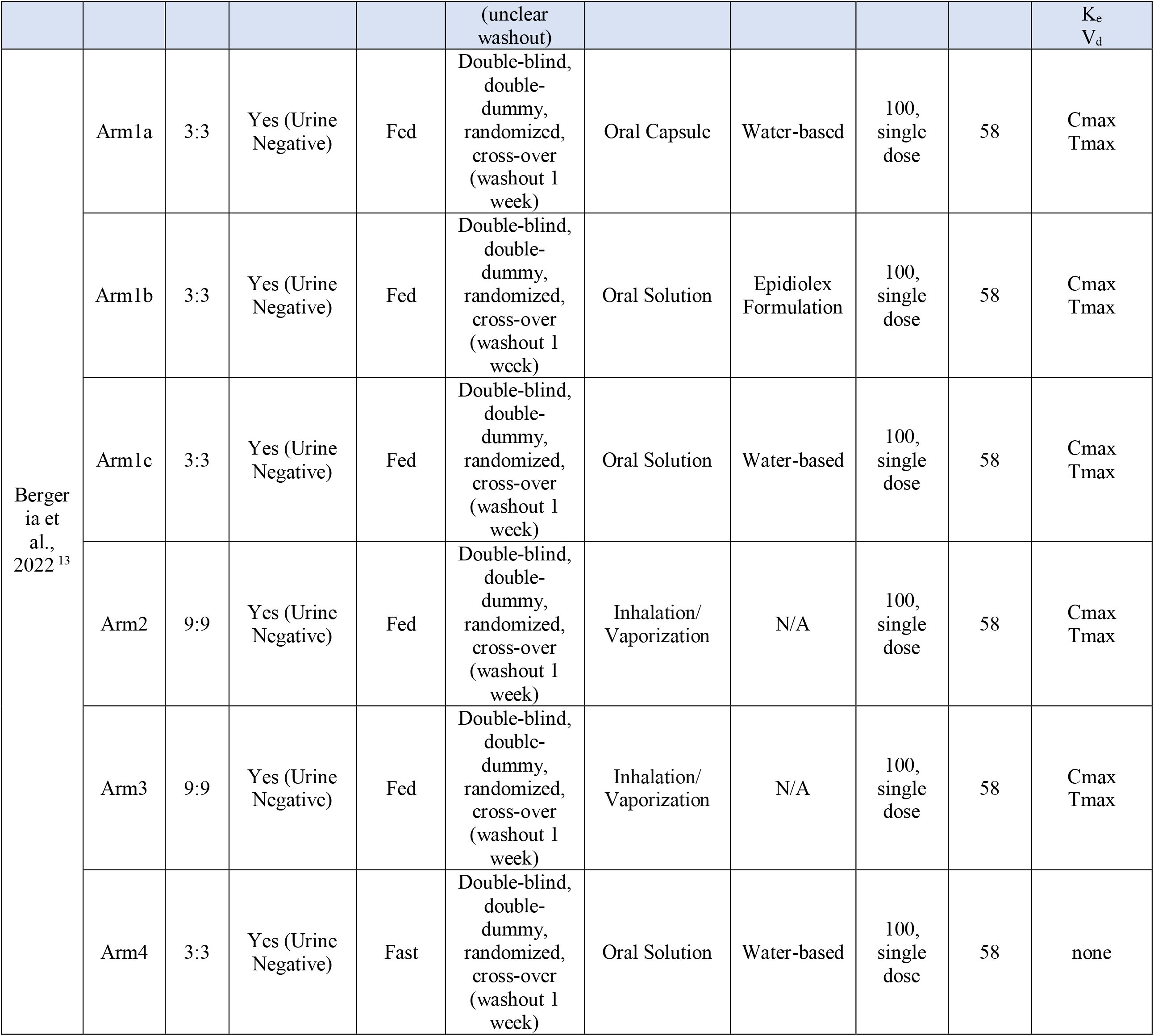
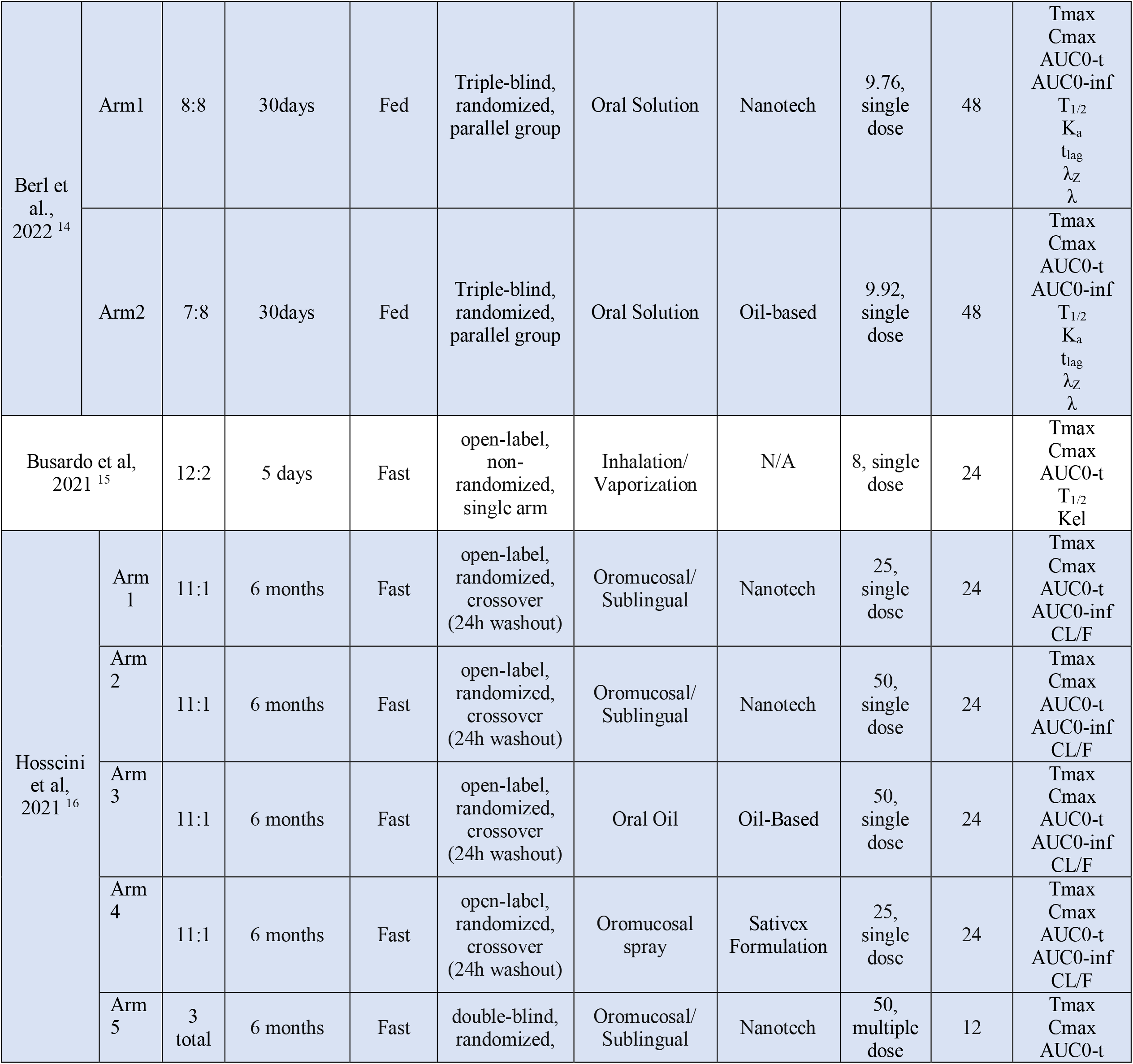
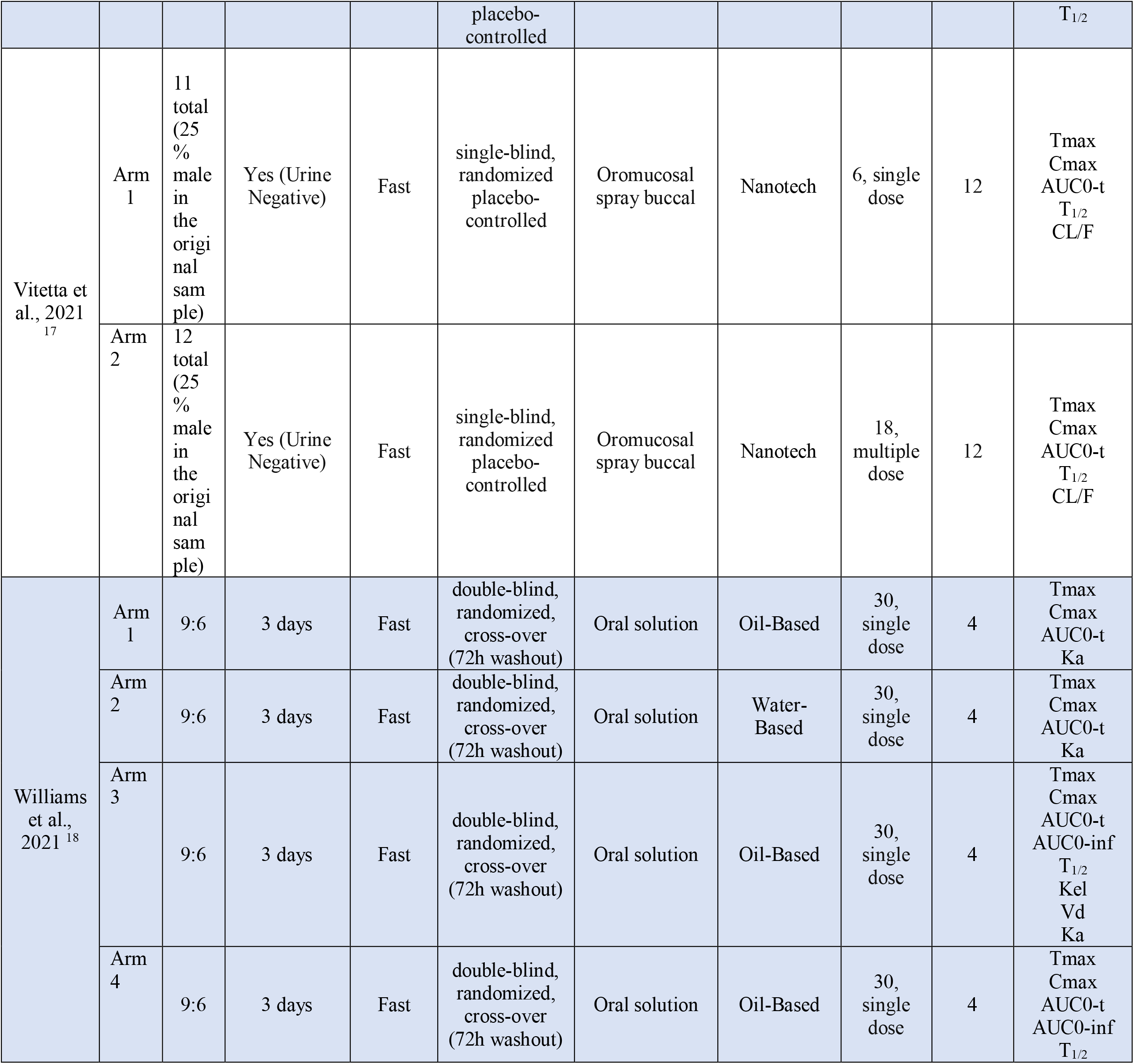
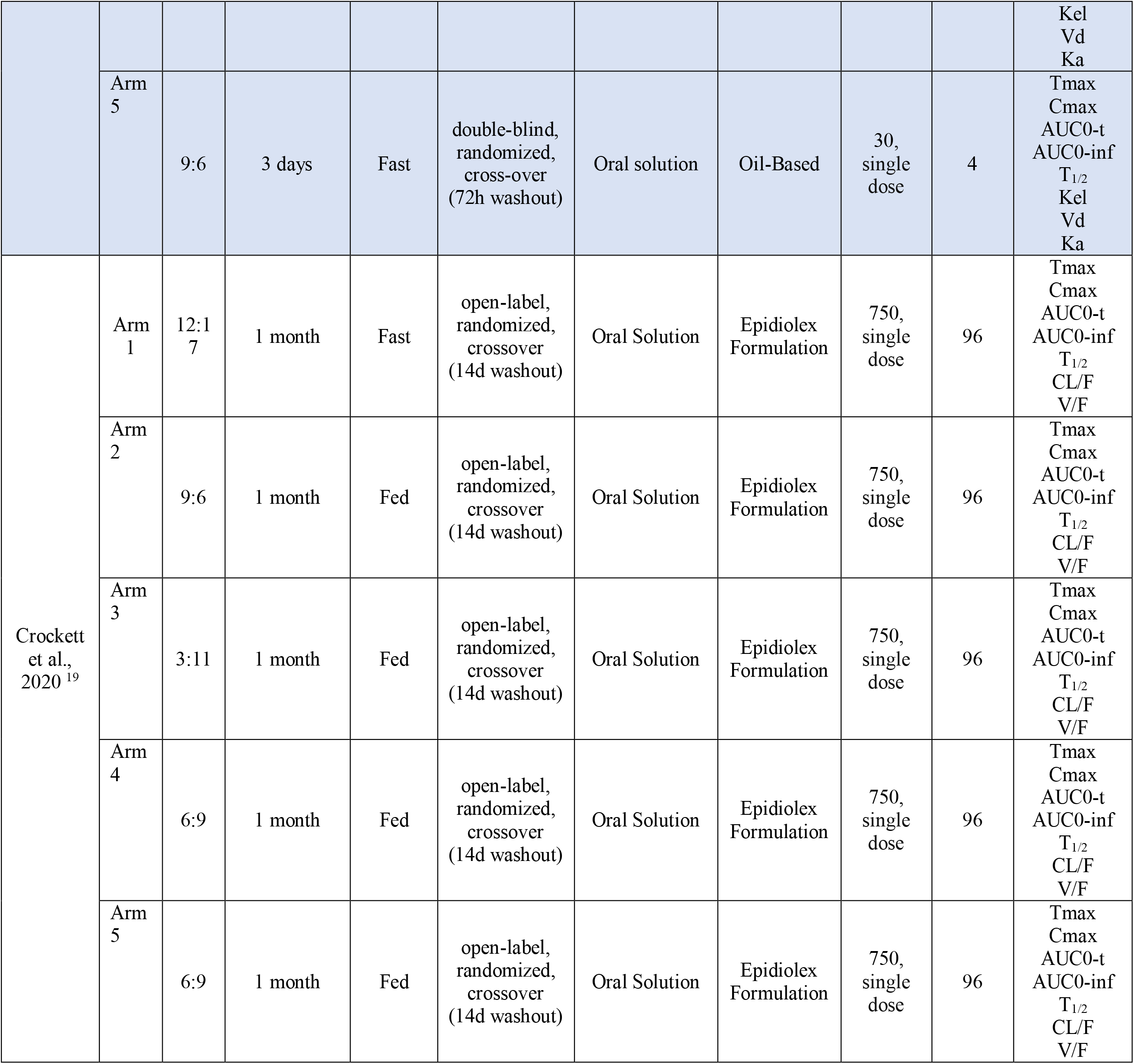
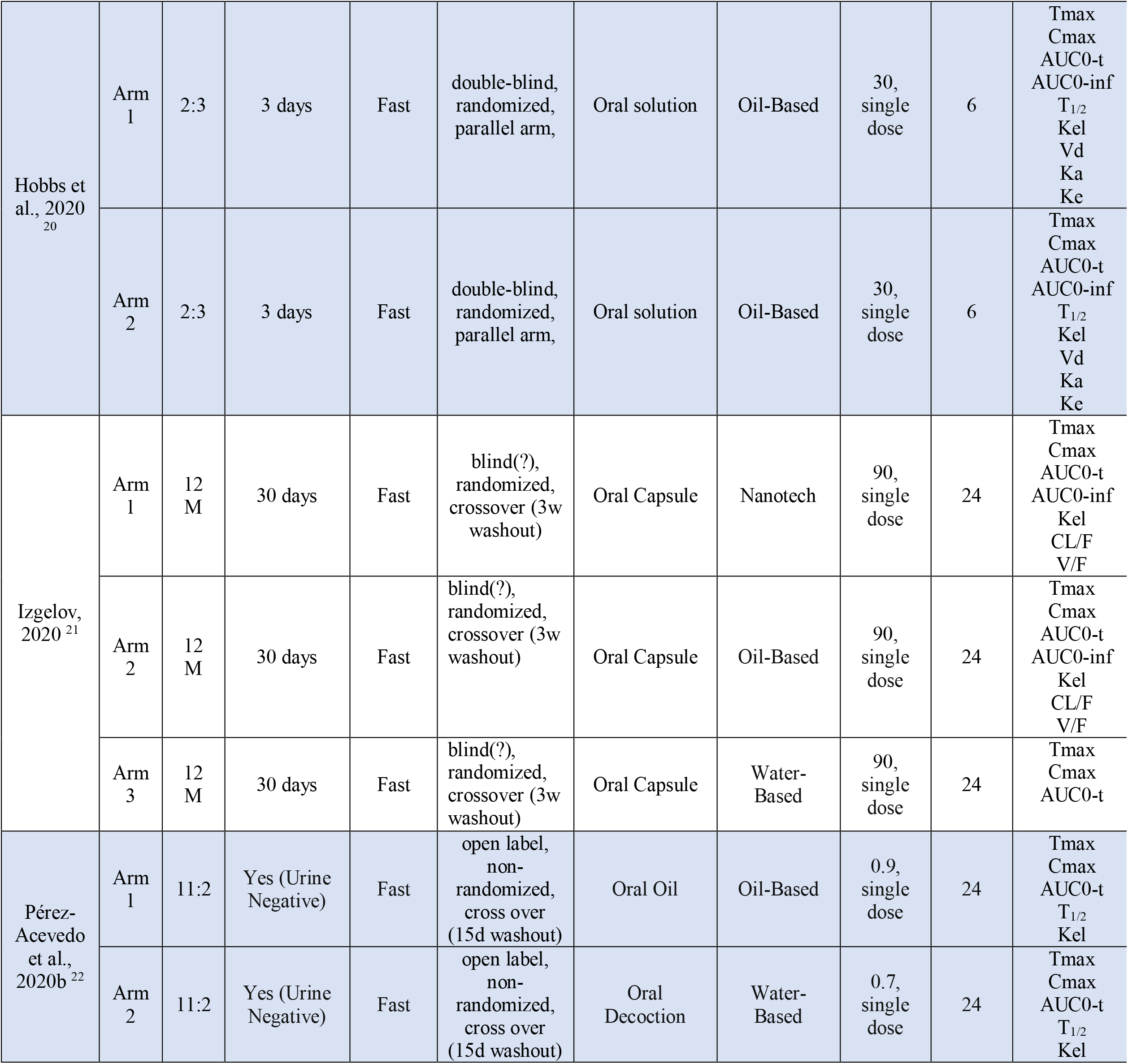
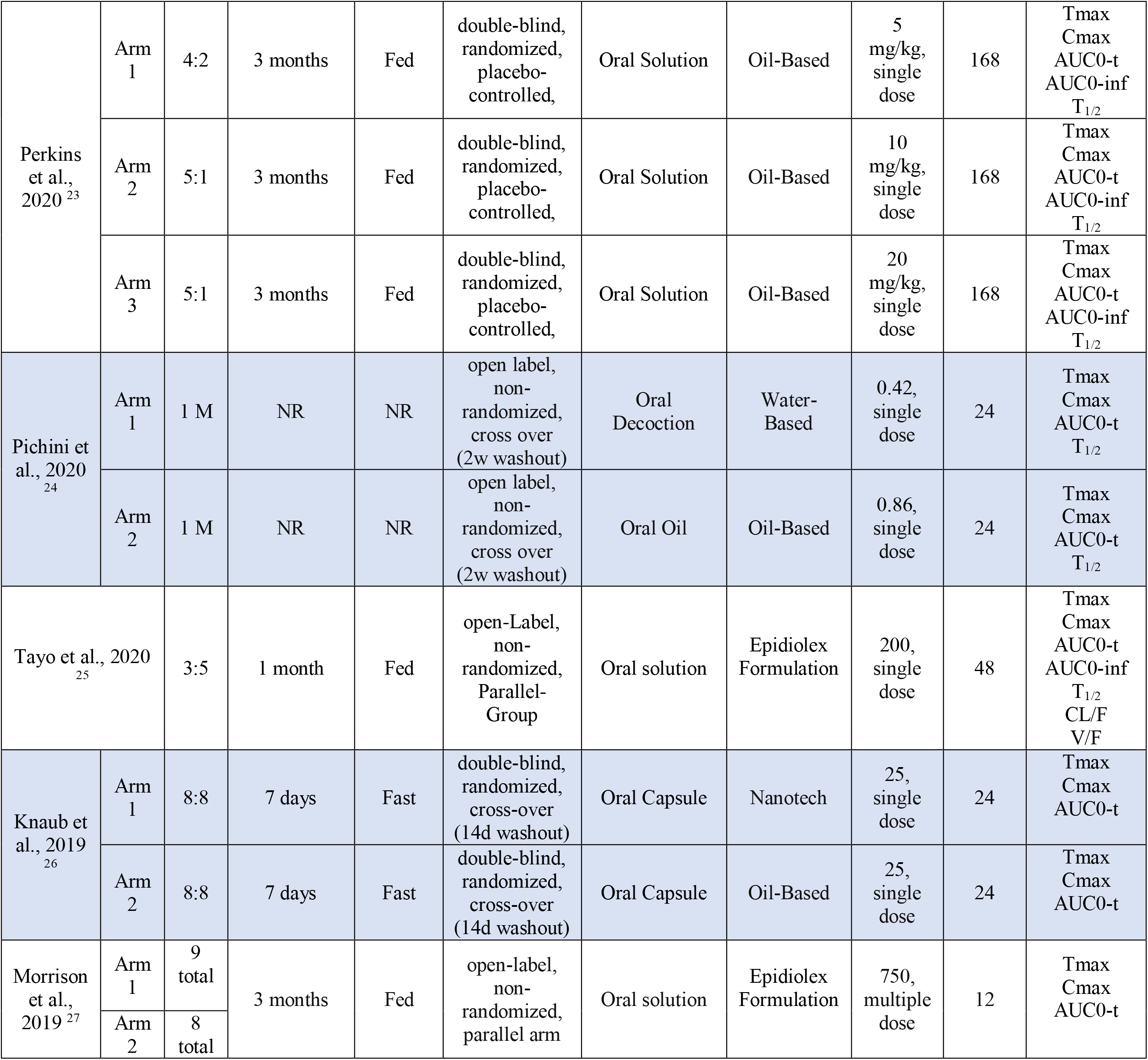
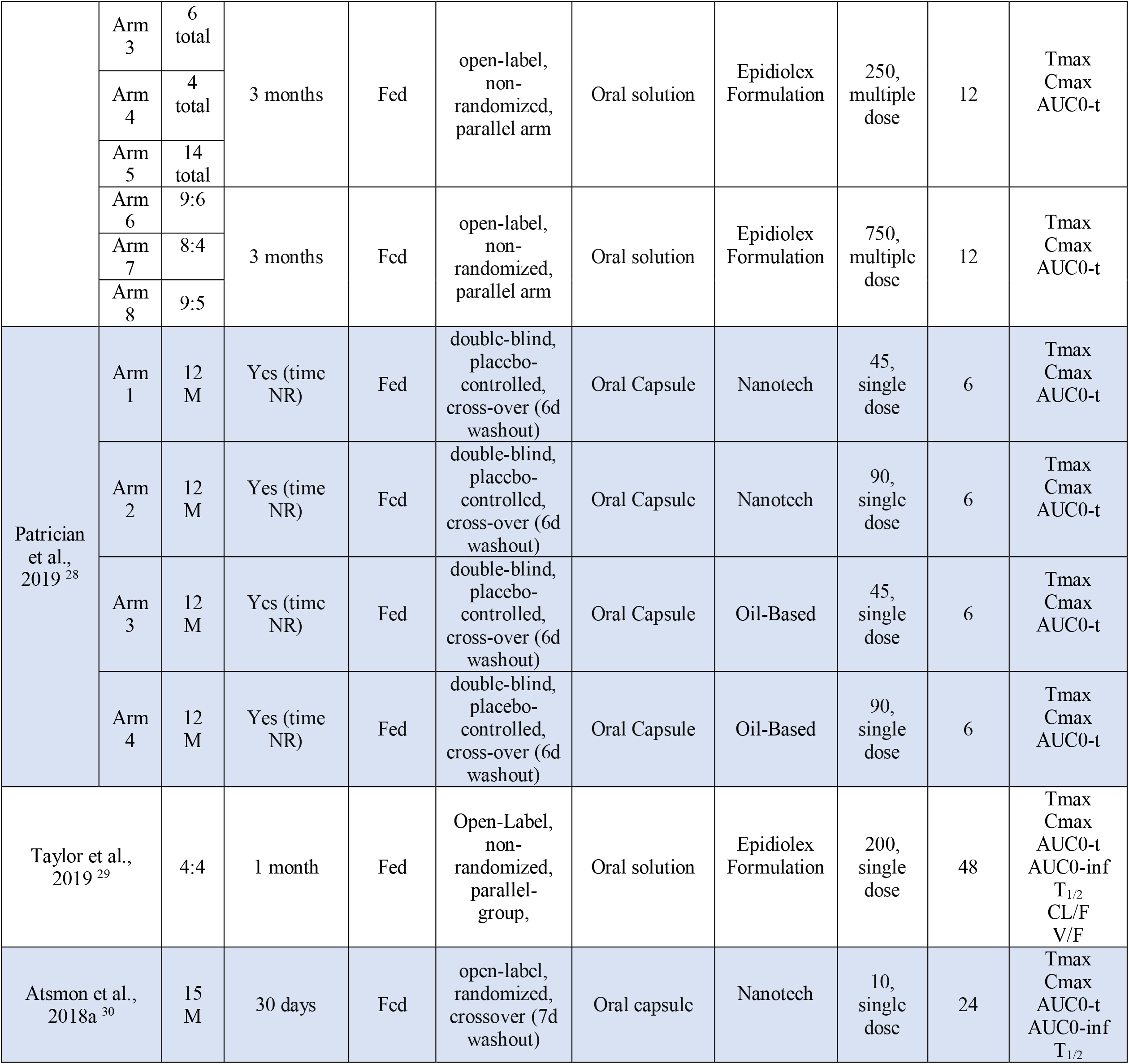
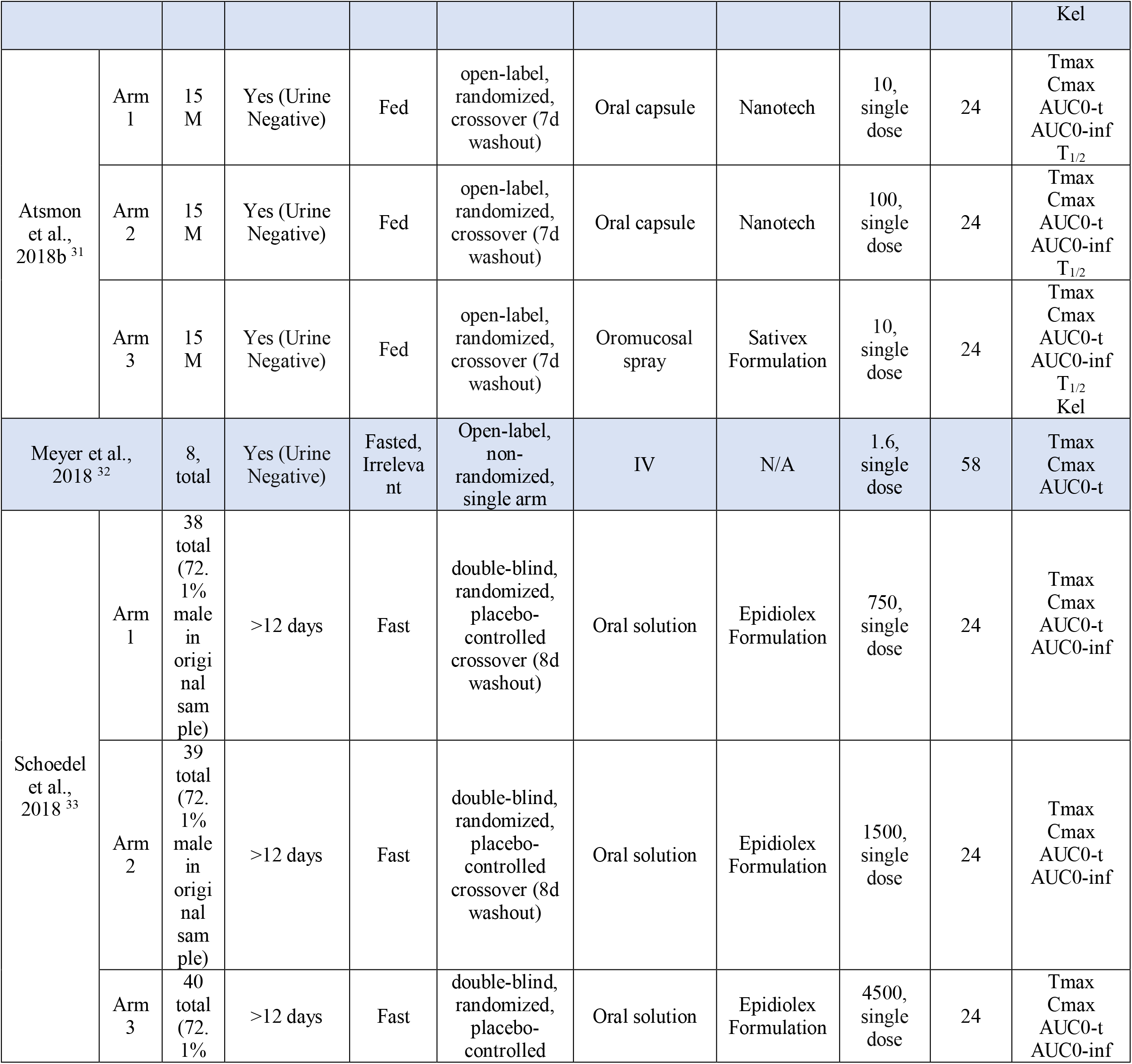
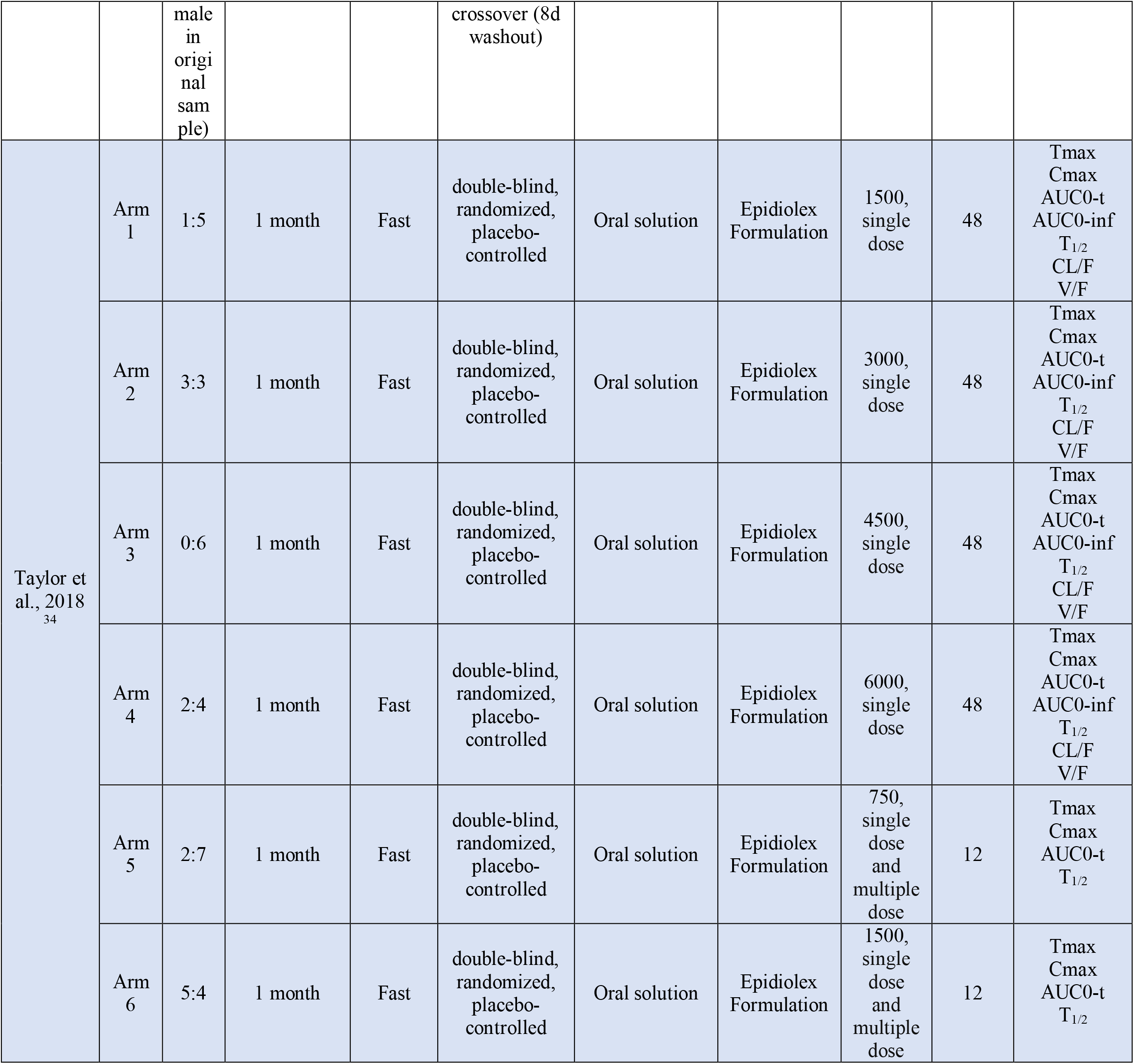
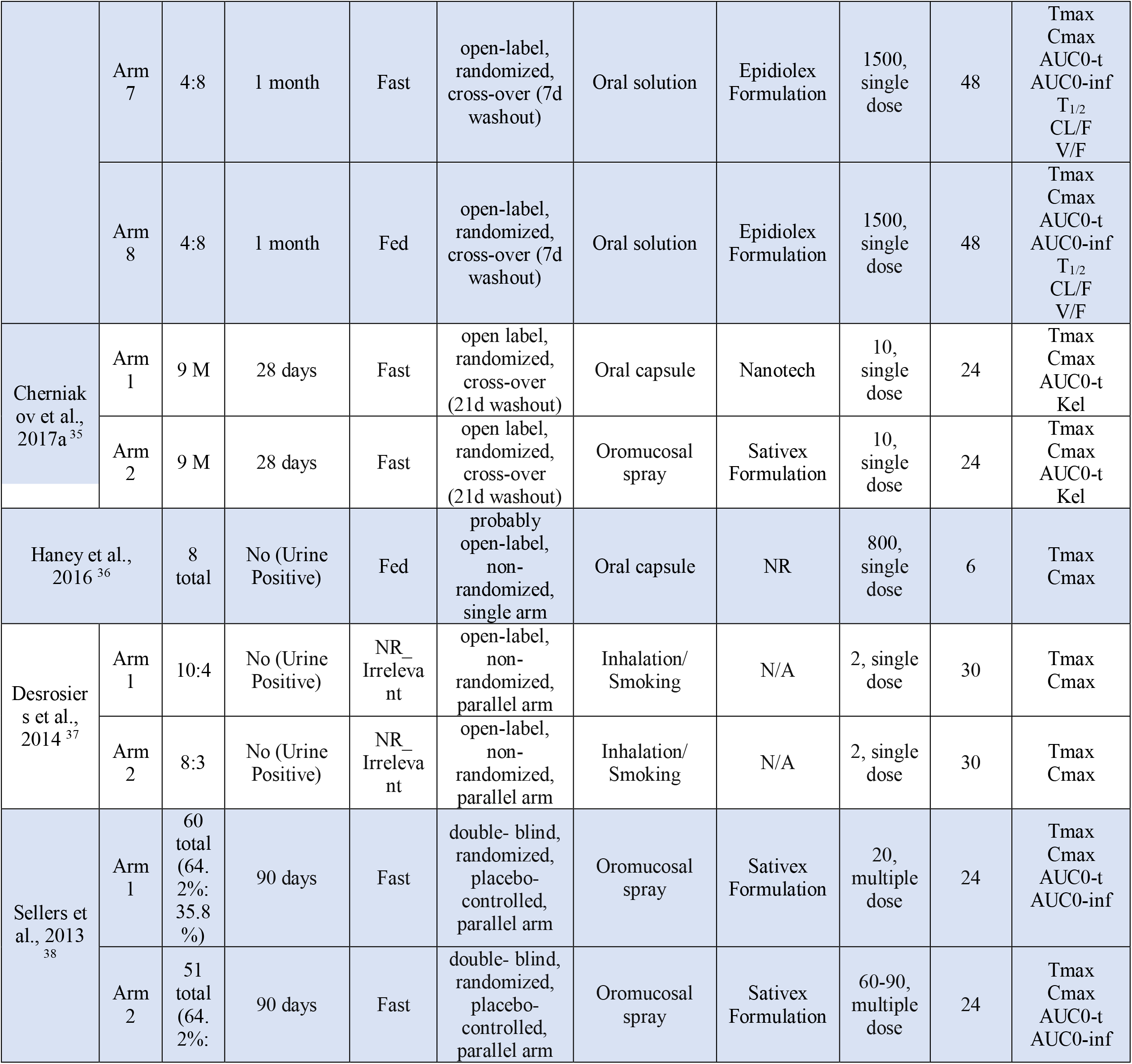
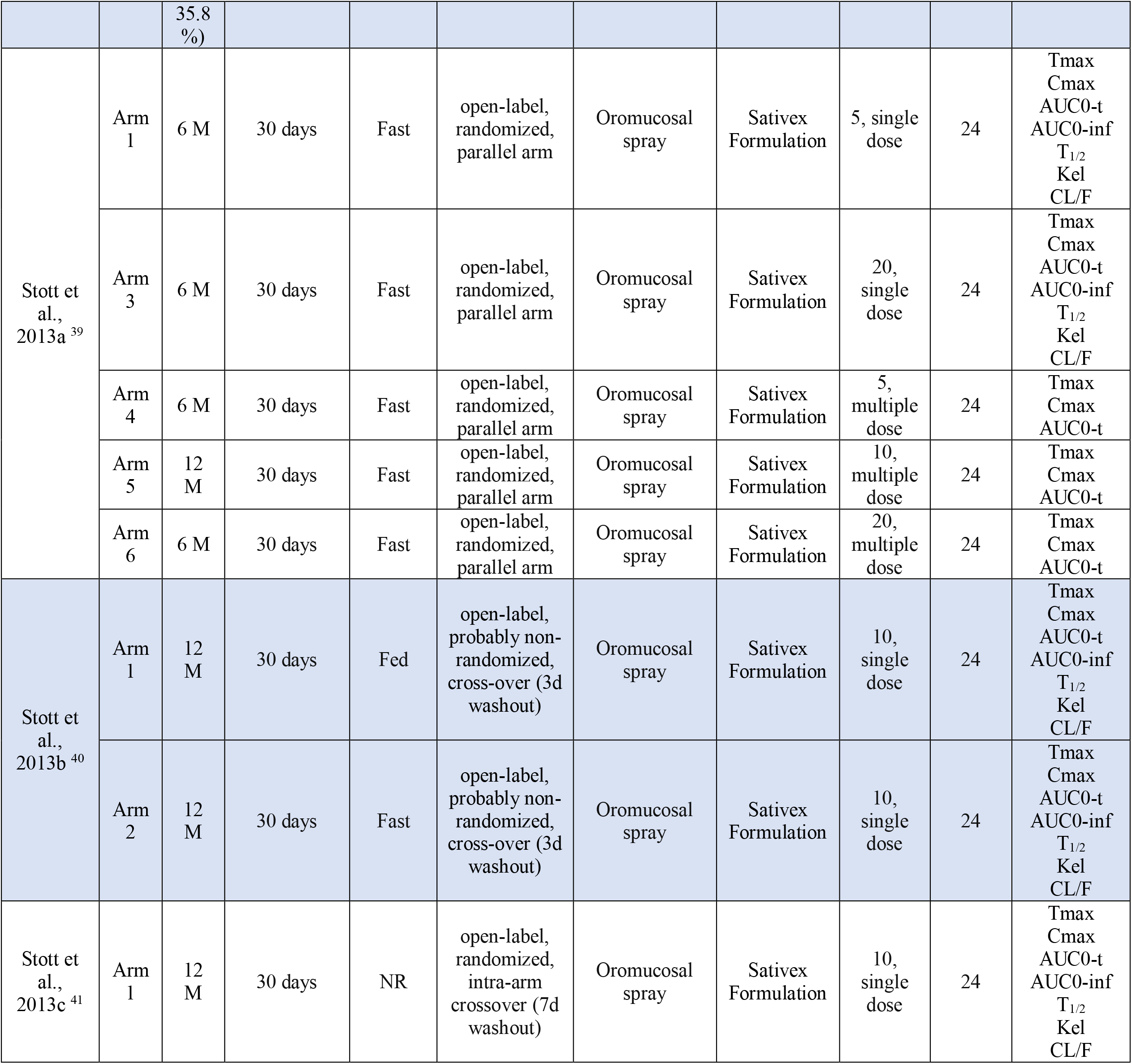
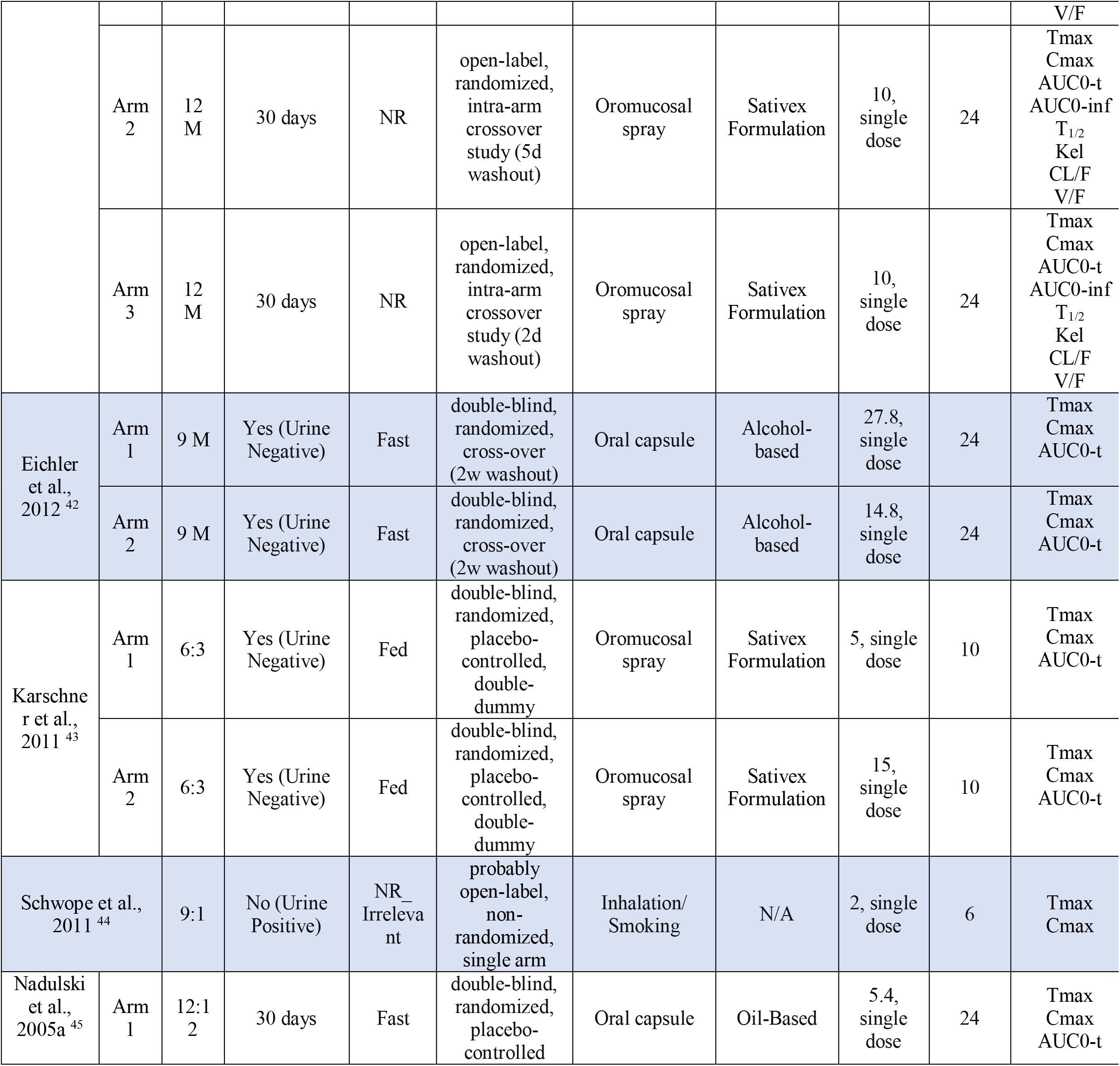
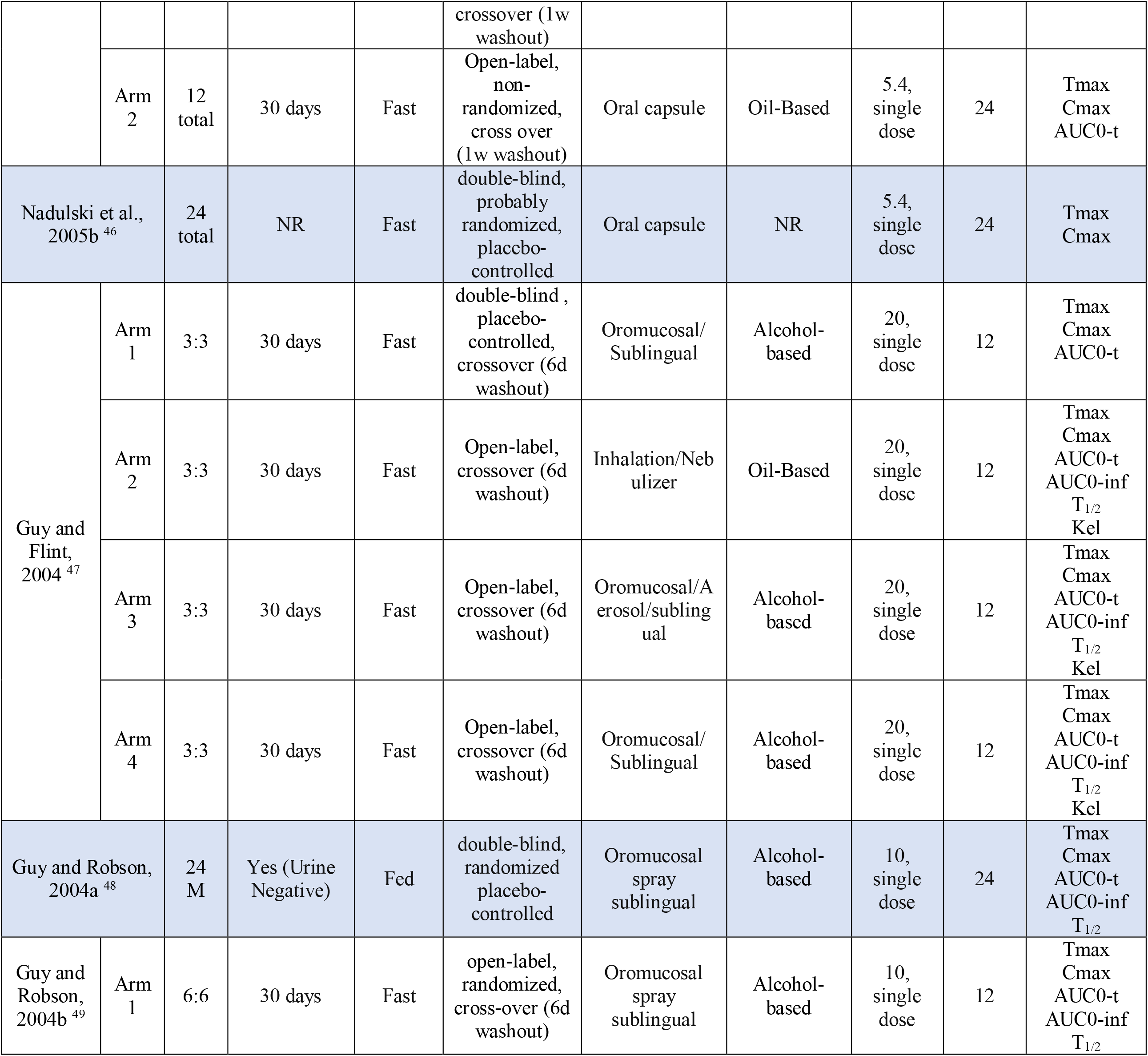
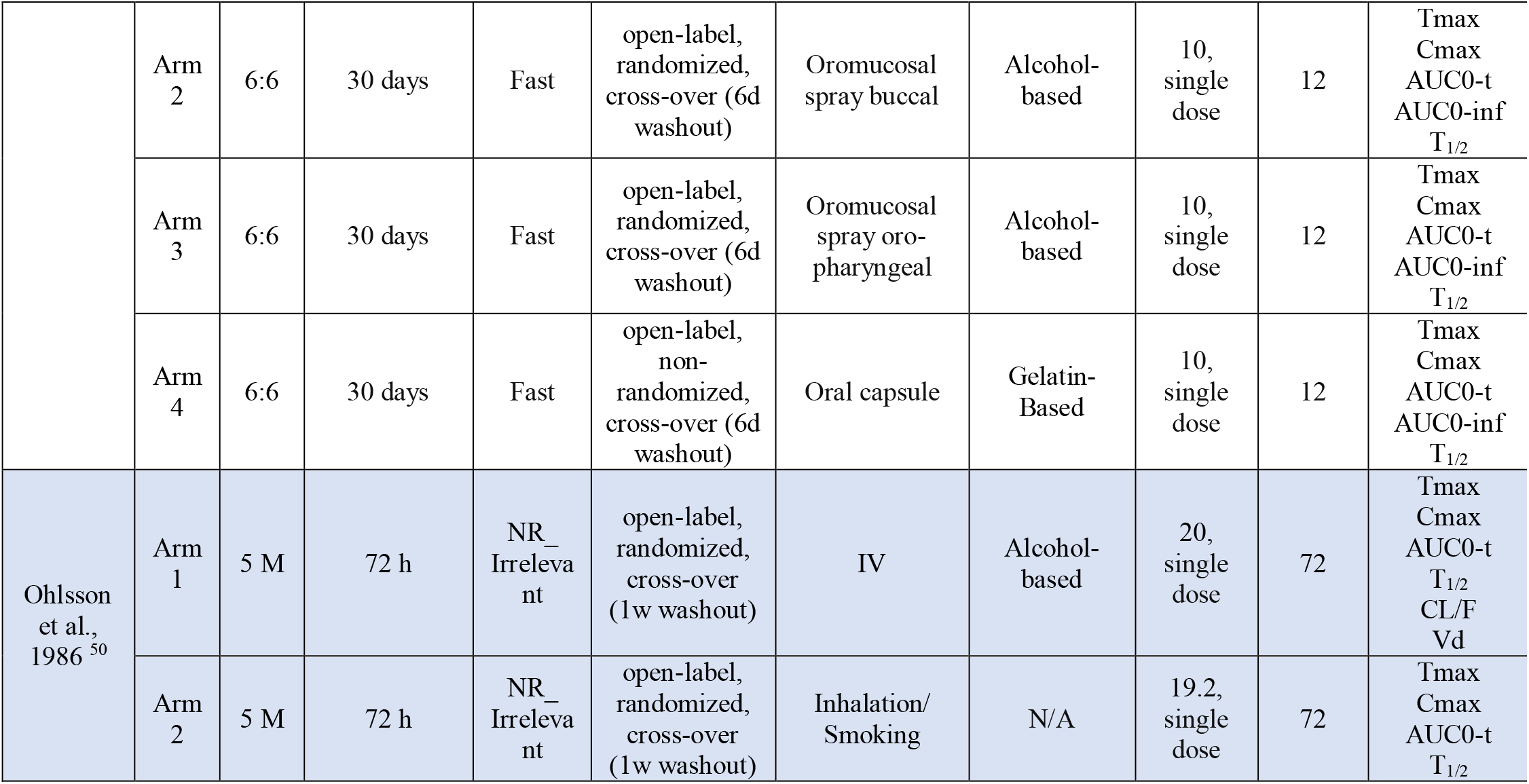
Summary of pharmacokinetic studies of cannabidiol

### Statistical analyses

To make between-study comparisons and meta-regression analysis feasible, the reported values for PK parameters went through two steps of conversion/estimation, using online calculators and SPSS v. 28 (IBM Statistics, USA). In the first step, values were converted into a unified form consisting of hours for Tmax and T_1/2_, ng/mL for Cmax, h*ng/mL for AUC0-t and AUC0-inf. In the second step, we converted the values for Tmax and T_1/2_ into arithmetic mean [standard deviation (SD)] format and all the values for Cmax, AUC0-t, and AUC0-inf in geometric mean [coefficient of variation (CV%)] format as the preferred method for reporting of PK parameters ^4^. This was carried out because Cmax, AUC0-t, and AUC0-inf are usually highly variable and skewed, so a geometric scale is preferrable. In contrast, Tmax and T_1/2_ are less variable and an arithmetic scale is an acceptable method which also would require less conversions in this review thus allowing for higher accuracy. For the linear meta-regression analysis, confidence intervals were estimated and log-transformed for every parameter’s values as necessary. Step 2 conversions are based on previously published methods and the latest guidelines ^9-11^, summarized and simplified in the supplementary material Section B to help with utilization in other PK reviews and replicability of this work. Missing data were addressed as follows: (1) if the mean or median was not reported for a specific PK parameter, it was considered missing and was not imputed; and (2) if no measure of distribution (e.g. SD or CV%) was reported for a PK parameter, but the mean or median was reported, the SD (for Tmax and T_1/2_) or CV% (for Cmax, AUC0-t, AUC0-inf) was imputed using the largest SD or CV% value available for that parameter among all single-dose trial arms.

Comprehensive Meta-Analysis v. 3 (Biostat Inc., USA) software was used for random-effects multivariable meta-regression analysis, with PK parameters as dependent outcomes and route of administration, CBD dose, diet status, CBD formulation, female ratio, and duration of PK session as independent predictors. Details of considerations for modeling are provided in the supplementary material Section C. Overall, three groups of models were built for each PK parameter across single-dose trial arms: (1) models including all the three routes of CBD administration (inhalation, oromucosal, and oral); (2) models including only oromucosal and oral trial arms; and (3) models including only oral CBD trial arms. For sensitivity analysis, models were conducted once with all the single-dose trial arms irrespective of the quality rating and then a second time only including trial arms with a “Good” or “Fair” quality rating. For each model, the number of included arms was reported alongside the R-squared value to measure model fit, indicating the amount of variability in the data that the model could explain. For each variable in the model, the significance level (alpha=0.05) and the positive or negative sign of the regression coefficient were reported to indicate a positive or negative association, respectively. Since models were based on log-transformed data, the net regression coefficient values were not interpretable in terms of effect size, and thus only their positive/negative sign was reported.

## Results

### Study selection and overview

After processing all the retrieved records, 39 studies comprising 112 trial arms were included in the narrative synthesis ^12-38^,^39-50^ out of which 35 studies comprising 92 arms were used for quantitative comparisons and regression analysis (Figure 1). Out of the 20 trial arms that were excluded from the analysis, but included in the narrative synthesis, 15 trial arms administered multiple doses of CBD (Table 1), two arms had only one participant ^24^, two arms were intravenous administration of CBD ^32,50^, and PK values was not reported for one trial arm ^13^.

**Figure 1.**
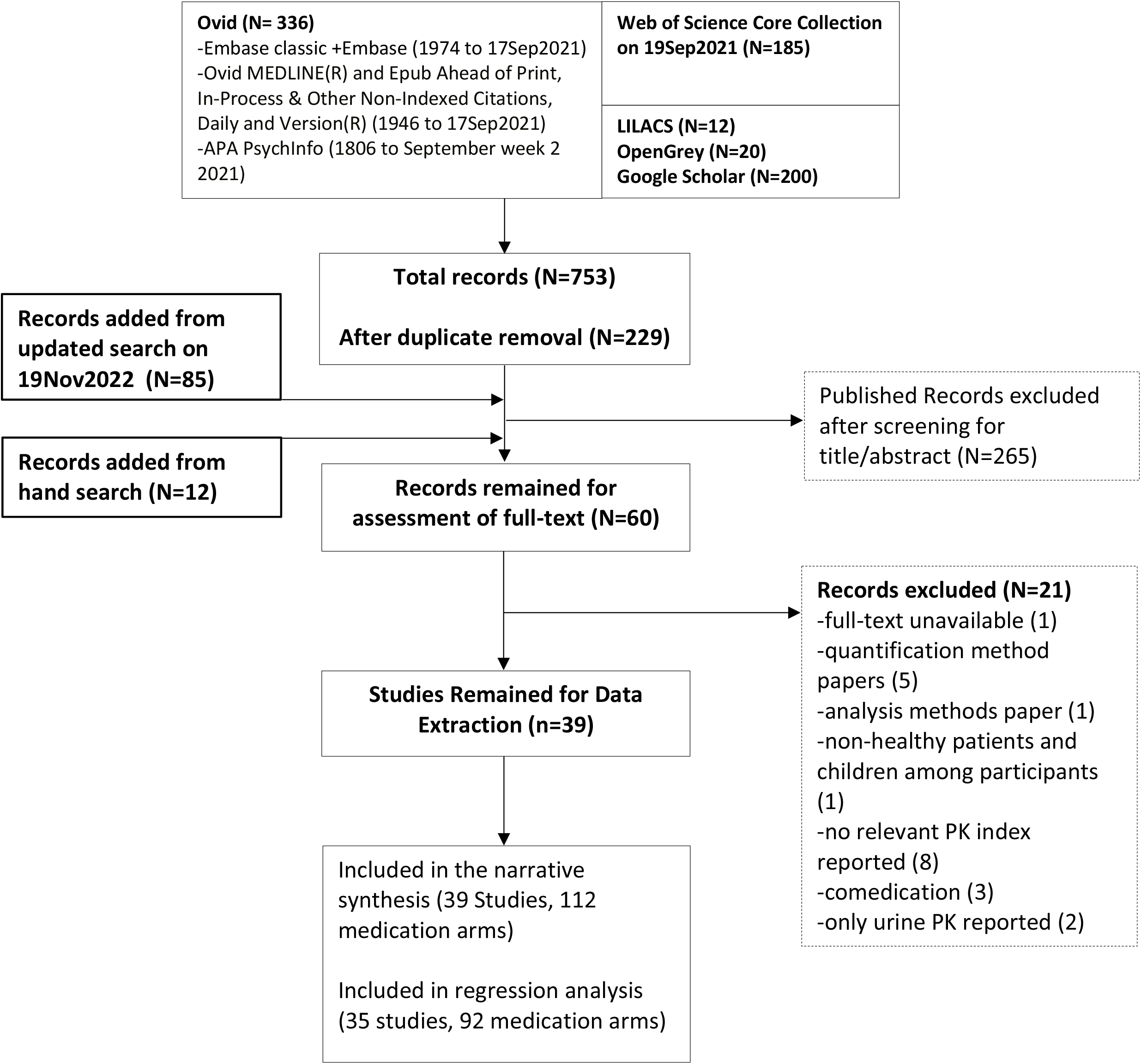
PRISMA flow diagram

### Quality assessment

Twenty-six trial arms were rated as “Good,” 70 as “Fair,” and 16 as “Poor” based on 12 criteria (Q1-Q12) (Table 2). Most studies clearly stated the study question (Q1), described eligibility criteria (Q2) and had a representative sample in terms of inclusion/exclusion criteria (Q3). At the same time, they mostly failed to report whether all the eligible potential participants were included (Q4), and most had either moderate or low sample sizes (Q5), which was an essential consideration for rating. The intervention was well described in most studies (Q6), and PK outcomes were well-defined and consistently assessed (Q7). The majority of trial arms were non-double-blind (Q8). Subject loss at follow-up (Q9) was reported in most studies. Statistical methods primarily measured before-after changes (Q10), for multiple times (Q11), except in some cases where it was not clearly stated. All studies were conducted at the individual participant level with no group interventions applicable to any of the studies (Q12).

**Table 2.**
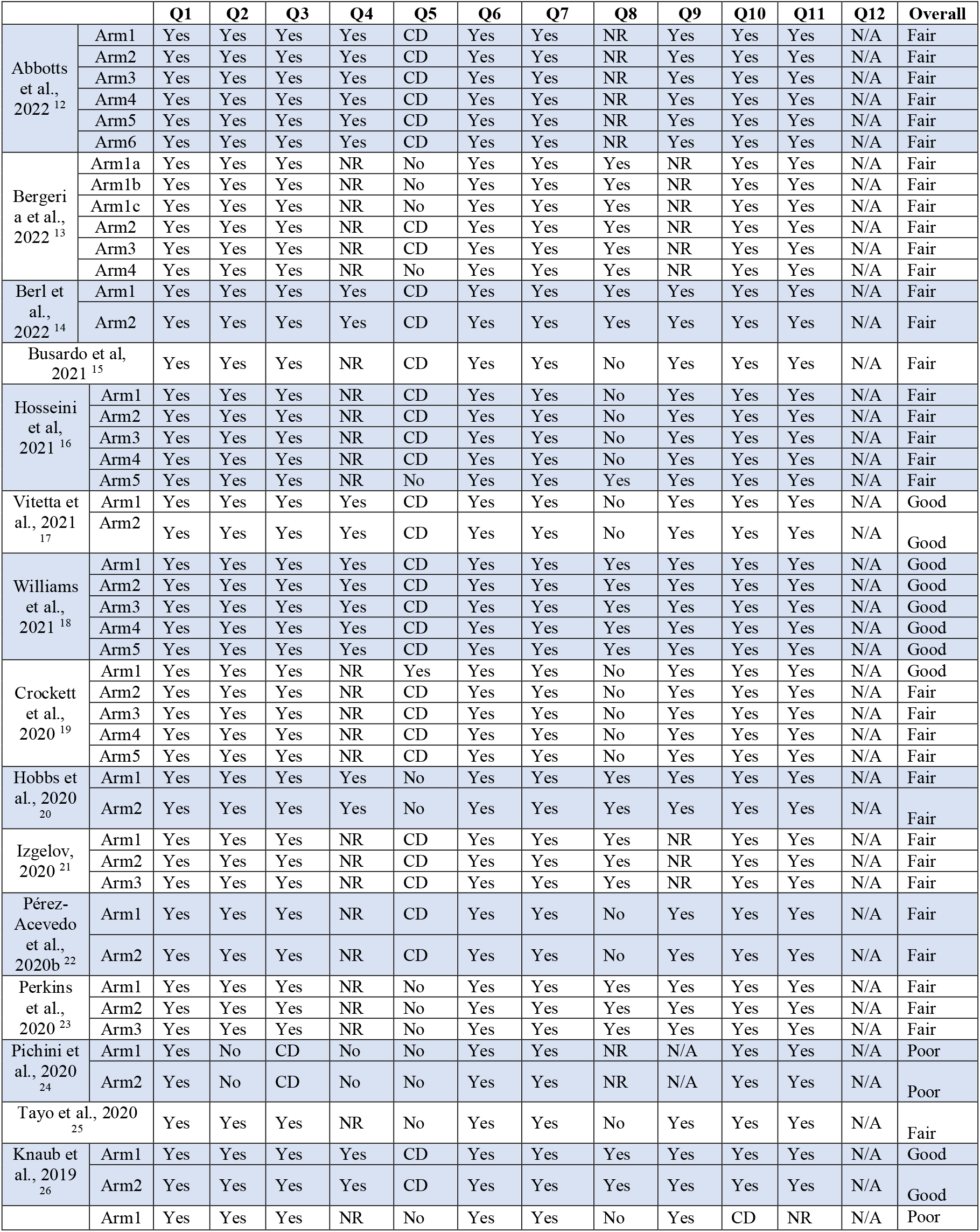
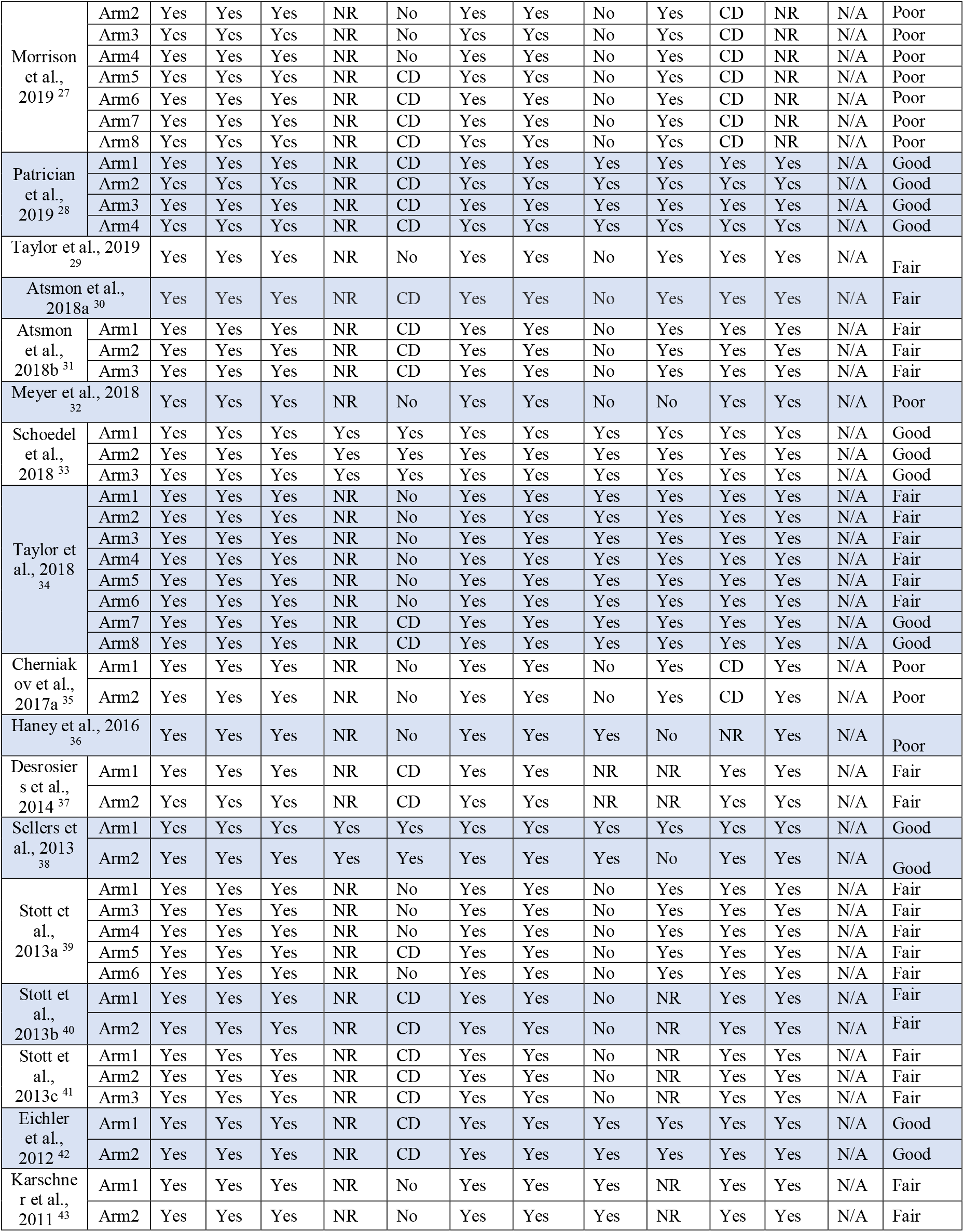
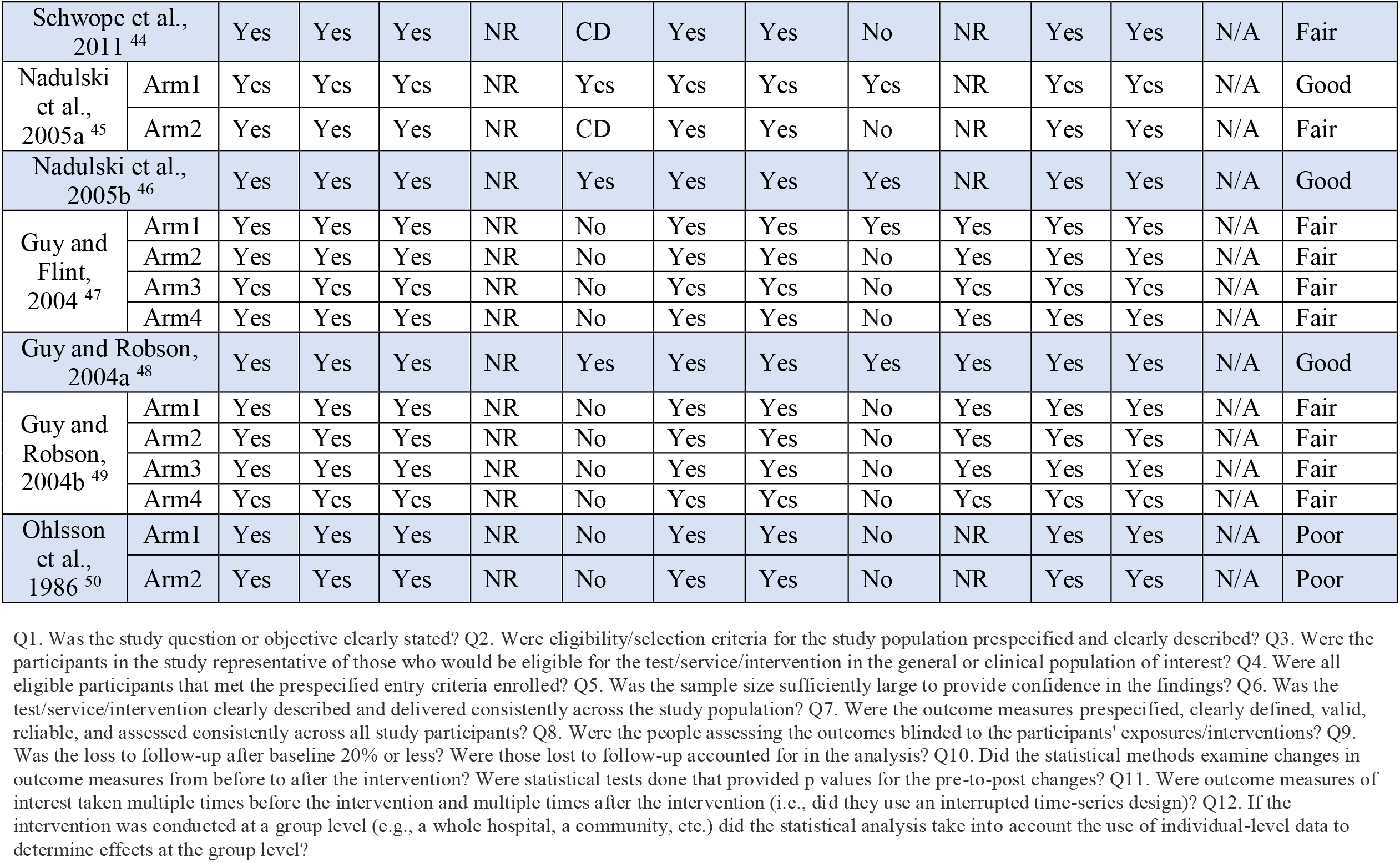
Quality assessment of pharmacokinetic studies of cannabidiol

### Narrative synthesis

Thirty studies had more than one treatment arm, out of which 22 studies, comprising 69 arms, had cross-over designs with washout periods ranging between 24h to 21days (Table 1).

Seventeen studies, comprising 42 arms, had at least one double-blind arm. Sixty-six trial arms had either only male participants or a higher number of males than females, while the sex ratio was either equal to one or favored females in 36 arms. Participants’ sex was not clearly reported for 10 arms. Participants were abstinent from cannabis before study initiation in 105 arms and had fasted before CBD administration in 63 arms. Eight arms used inhalation as the route of administration, 29 oromucosal, 73 oral, and 2 intravenous. A variety of formulations were used consisting of nanotech (n=14), oil-based (n=21), alcohol-based (n=10), and water-based (n=12), alongside Sativex (n=17) and Epidiolex (n=22) formulations. For single-dose studies, CBD doses ranged between 2-100mg in inhalation, 5-50mg in oromucosal, and 0.42-6000mg in oral administration (Table 1). The duration of the PK session was between 4-164 h. All trial arms reported Tmax and Cmax except one that did not report any values, 96 reported AUC0-t, 59 reported AUC0-inf, and 60 reported T_1/2_. Only 22 treatment arms from four studies reported Cmax and AUC in geometric scale (Supplement Table1 and Supplement Showcase). At least one PK parameter from each and all treatment arms needed to go through second step conversions to geometric values in order to conform to the reporting format in Supplement Table 1.

### Quantitative synthesis

#### Reported PK Values

Given the large variability of the doses studied, Table 3 provides a summary of the PK parameters for doses consisting of more than 2 trials. Suppl Table 1 presents the PK parameters from all single-dose trial arms. Figure 2 demonstrates the PK parameters in order of increasing CBD dose for all the trial arms. Suppl Figure 1 is a magnified version of Figure 2 for CBD doses equal or less than 100mg. Among single-dose trial arms, the arithmetic mean Tmax ranged between 0.00-0.60 h for inhalation, 1.00-5.01 h for oromucosal, and 0.59-10.45 h for oral administration (Suppl Table1, Figure 2). Geometric mean Cmax ranged between 0.42-120.77 ng/mL for inhalation, 0.38-12.90 ng/mL for oromucosal, and 0.22-1628 ng/mL for oral administration. Geometric mean AUC0-t ranged between 6.18-76.77 h*ng/mL for inhalation, 0.69-61.64 h*ng/mL for oromucosal, and 0.47-9390.94 h*ng/mL for oral administration. The geometric mean AUC0-inf was 9.03 for the only inhalation study that reported this parameter, ranging between 1.59-70.98 h*ng/mL for oromucosal and 3.32-8669 h*ng/mL for oral administration. The arithmetic mean T_1/2_ ranged between 1.10-31.00 h for inhalation, 1.44-10.86 h for oromucosal, and 1.09-70.3 h for oral administration.

**Table 3.**
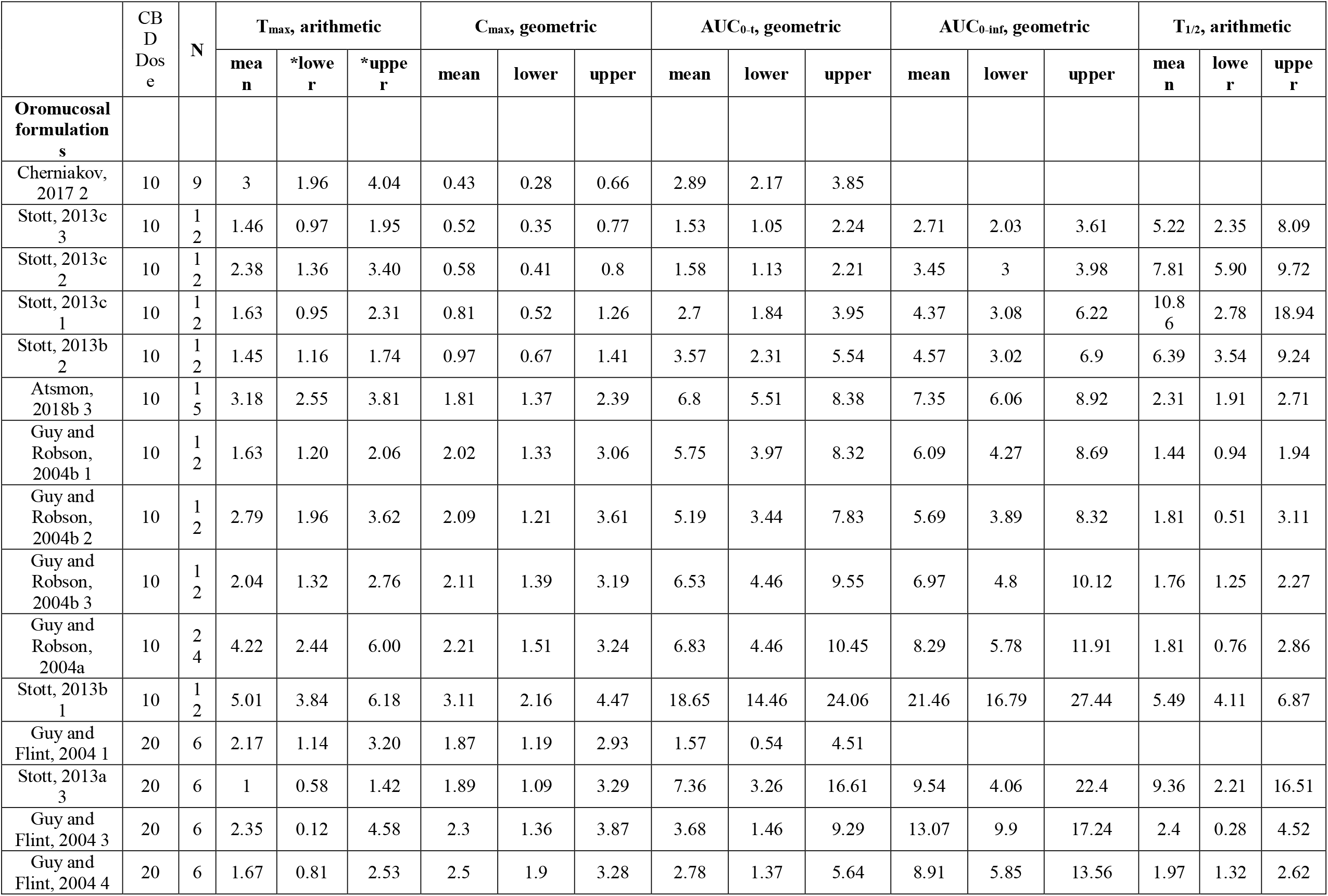
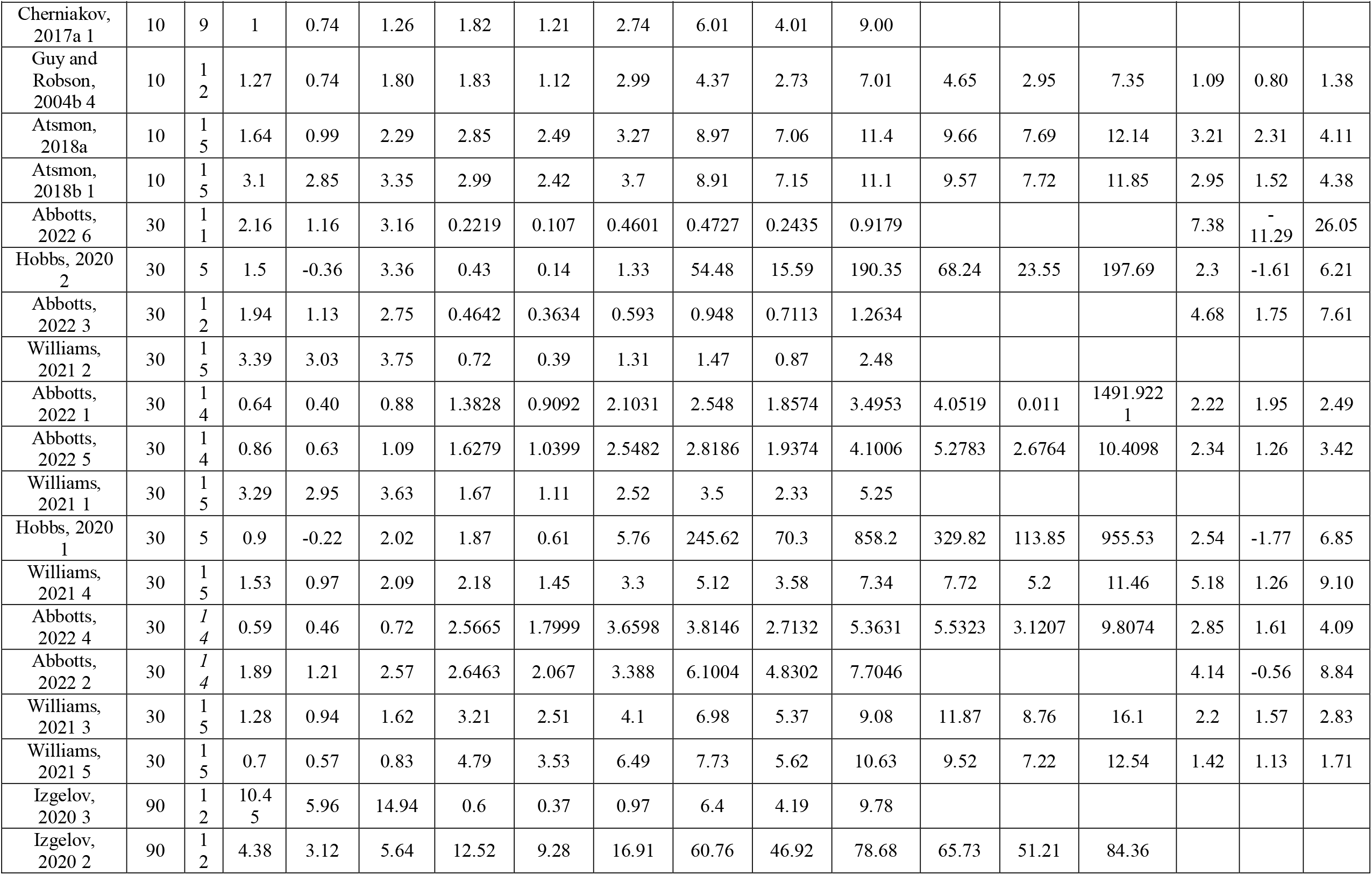
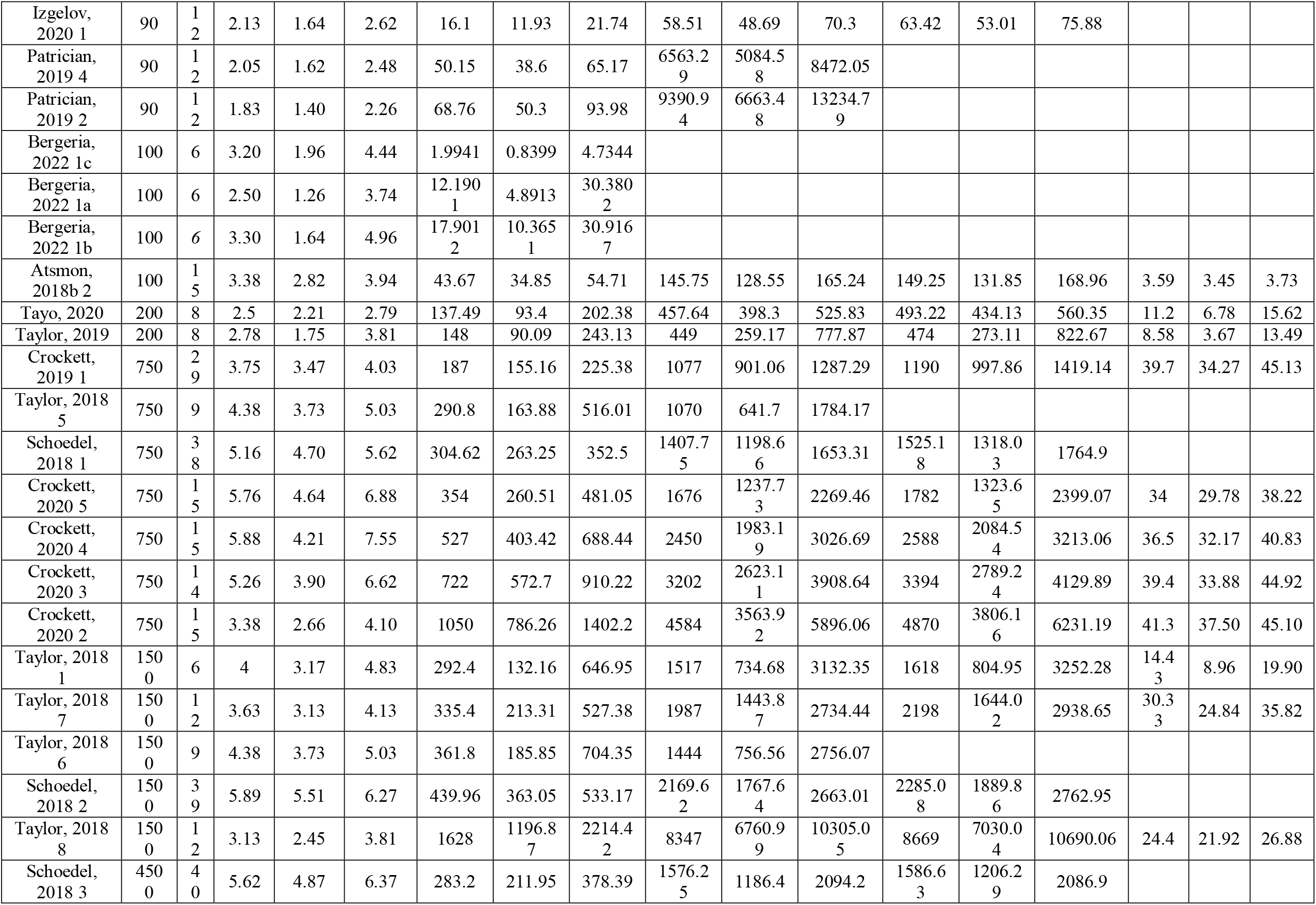
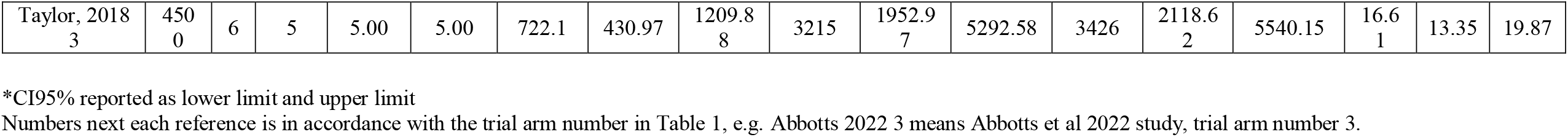
Pharmacokinetic parameters of cannabidiol in single dose studies in accordance with increasing CBD dose

**Figure 2.**
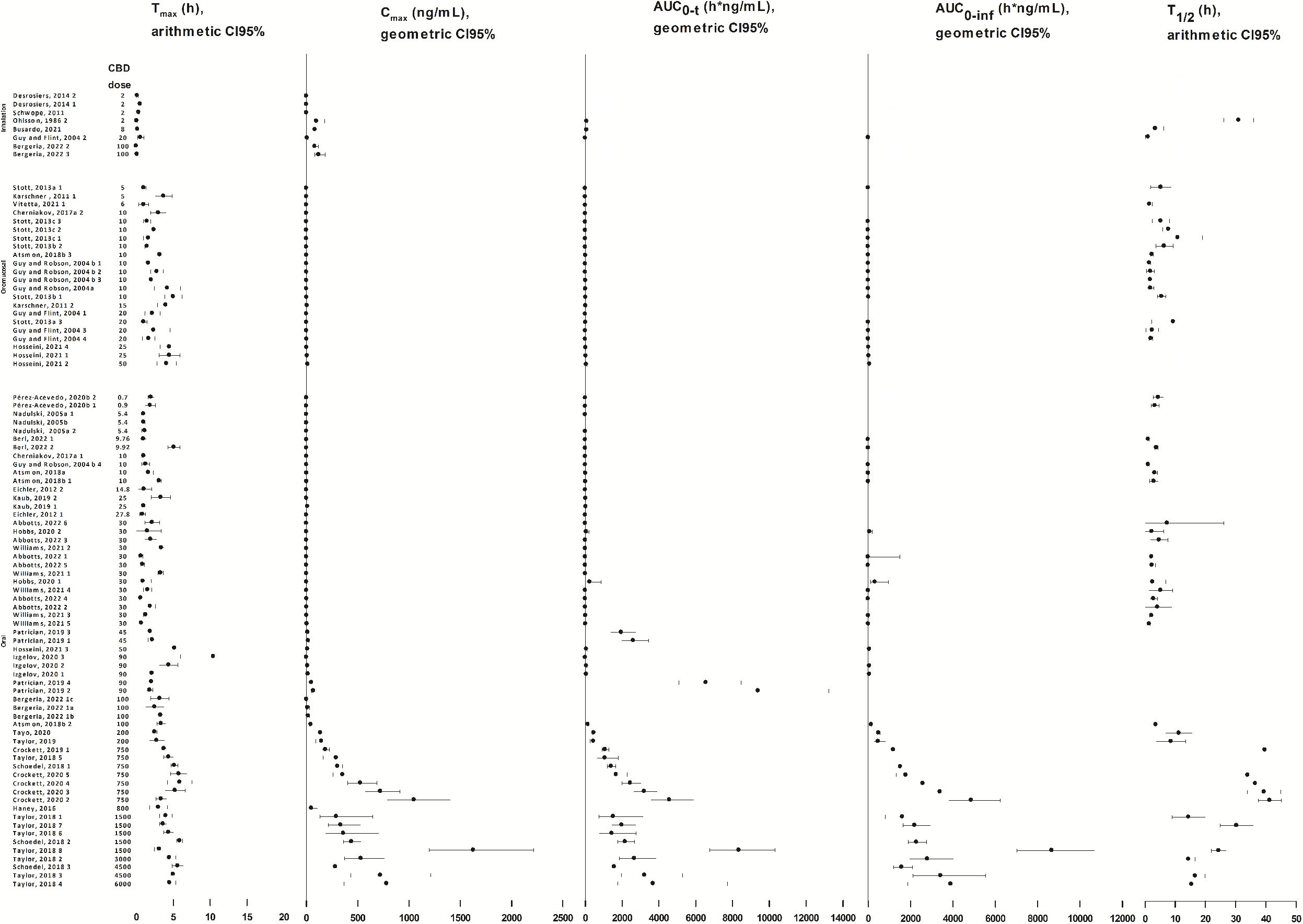
Pharmacokinetic parameters of single dose cannabidiol studies in increasing order of CBD dose.

#### Meta-regression analysis

Overall, out of a total number of 92 trial arms that were included in analysis, 88 were used in regression models for Tmax, 86 for Cmax, 78 for AUC0-t, and 53 for AUC0-inf and T_1/2_. The amount of variability in the data that the models could explain (R^2^) ranged between 0-83% for Tmax, indicating a very low to high model fit, 41-49% for Cmax, indicating a moderate fit, 44-52% for AUC0-t indicating a moderate fit, 35-70% for AUC0-inf indicating a low to high fit, and 84-87% for T_1/2_ indicating a high fit. Removing the “Poor” quality trial arms from the models did not result in a noticeable change in model fit or a change in the significance of variables.

Higher CBD dose was consistently associated with higher Cmax, AUC0-t, and AUC0-inf across all models (Table 4; Figure 2; Suppl Figure 1). Compared to oral administration as a reference, inhalation was associated with lower Tmax in a poorly-fitted model (Table 4, Model# 1,2), but did not show any significant difference for Cmax (Model# 7,8), AUC0-t (Model# 13,14), or AUC0-inf (Model#19). In addition, compared to oral administration as a reference, oromucosal administration was associated with lower Cmax (Model# 7-10), AUC0-t (Model# 13-16), and AUC0-inf (Model#19,20), but there was no significant difference for Tmax (Model# 1-4). Compared to the Epidiolex formulation as a reference, nanotech and oil-based formulations were associated with a lower Tmax (Model#5,6). Fed status was associated with higher Cmax (Model# 9-12) and AUC0-t values for both oromucosal and oral administration models (Model#15-18) compared with the fasting status. No significant association of fed status was observed with either Tmax (Model# 3-6) or AUC0-inf (Model#20-21). A higher ratio of female participants in the sample was associated with lower Tmax (Model# 5,6) and higher T_1/2_ (Model# 25) only among oral administration arms. A higher ratio of female participants was also associated with higher Cmax (Model# 9-12) in all models. There was no significant association of female/total ratio with AUC0-t (Model#13-18) or AUC0-inf (Model# 19-21). Finally, longer study duration was associated with higher AUC0-t only in the regression models that included all routes of administration (Model# 13,14), and higher AUC0-inf and T_1/2_ in all the regression models (Model# 19-25).

**Table 4.** Meta-Regression models of pharmacokinetic parameters in single-dose cannabidiol studies

| Parameter | Model # | Route of administration | Number of arms | Model fit R<sup>2</sup> | CBD Dose | *Route: |  | *Formulation |  | *Diet: Fed | †Female / Total Ratio | Duration |
| --- | --- | --- | --- | --- | --- | --- | --- | --- | --- | --- | --- | --- |
|  |  |  |  |  |  | Inhalation | Oromucosal | Nanotechnology | Oil-based |  |  |  |
| T<sub>max</sub> | 1 | All routes | 88 | 0 |  | (-) <0.001 | (-) 0.936 |  |  |  | (+) 0.240 |  |
|  | 2 | All routes-fair/good | 85 |  |  | (-) <0.001 | (-) 0.831 |  |  |  | (+) 0.331 |  |
|  | 3 | Oral and oromucosal | 78 | 0.35 |  |  | (+) 0.828 |  |  | (+) 0.090 | (+) 0.166 |  |
|  | 4 | Oral and oromucosal-fair/good | 76 | 0.35 |  |  | (+) 0.946 |  |  | (+) 0.100 | (+) 0.186 |  |
|  | 5 | Oral | 49 | 0.83 |  |  |  | (-) <0.001 | (-) <0.001 | (+) 0.603 | (-) 0.007 |  |
|  | 6 | Oral-fair/good | 48 | 0.83 |  |  |  | (-) <0.001 | (-) <0.001 | (+) 0.821 | (-) 0.004 |  |
| C<sub>max</sub> | 7 | All routes | 86 | 0.41 | (+) <0.001 | (+) 0.406 | (-) 0.010 |  |  |  | (+) 0.011 |  |
|  | 8 | All routes-fair/good | 83 | 0.41 | (+) <0.001 | (+) 0.822 | (-) 0.014 |  |  |  | (+) 0.009 |  |
|  | 9 | Oral and oromucosal | 75 | 0.49 | (+) <0.001 |  | (-) 0.050 |  |  | (+) <0.001 | (+) 0.011 |  |
|  | 10 | Oral and oromucosal-fair/good | 73 | 0.48 | (+) <0.001 |  | (-) 0.067 |  |  | (+) <0.001 | (+) 0.015 |  |
|  | 11 | Oral | 56 | 0.47 | (+) <0.001 |  |  |  |  | (+) <0.001 | (+) 0.020 |  |
|  | 12 | Oral-fair/good | 55 | 0.47 | (+) <0.001 |  |  |  |  | (+) <0.001 | (+) 0.021 |  |
| AU C<sub>0-t</sub> | 13 | All routes | 78 | 0.44 | (+) <0.001 | (-) 0.747 | (-) <0.003 |  |  |  | (+) 0.583 | (+) 0.001 |
|  | 14 | All routes-fair/good | 75 | 0.44 | (+) 0.001 | (-) 0.887 | (-) <0.004 |  |  |  | (+) 0.723 | (+) 0.001 |
|  | 15 | Oral and oromucosal | 72 | 0.52 | (+) <0.001 |  | (-) <0.010 |  |  | (+) <0.001 | (+) 0.302 | (+) 0.094 |
|  | 16 | Oral and oromucosal-fair/good | 70 | 0.51 | (+) <0.001 |  | (-) <0.012 |  |  | (+) <0.001 | (+) 0.348 | (+) 0.094 |
|  | 17 | Oral | 53 | 0.49 | (+) <0.001 |  |  |  |  | (+) <0.001 | (+) 0.128 | (+) 0.510 |
|  | 18 | Oral-fair/good | 52 | 0.49 | (+) <0.001 |  |  |  |  | (+) <0.001 | (+) 0.143 | (+) 0.505 |
| AU C<sub>0-inf</sub> | 19 | All routes-fair/good | 51 | 0.36 | (+) 0.001 | (-) 0.463 | (-) 0.025 |  |  |  | (+) 0.201 | (+) <0.001 |
|  | 20 | Oral and oromucosal-fair/good | 47 | 0.35 | (+) 0.001 |  | (-) 0.054 |  |  | (-) 0.600 | (+) 0.350 | (+) <0.001 |
|  | 21 | Oral-fair/good | 33 | 0.70 | (+)<br>0.002 |  |  |  |  | (-)<br>0.531 | (+)<br>0.341 | (+)<br>0.001 |
| T<sub>1/2</sub> | 22 | All routes | 53 | 0.84 |  |  |  |  |  |  | (+)<br>0.493 | (+)<br><0.001 |
|  | 23 | All routes-fair/good | 52 | 0.86 |  |  |  |  |  |  | (+)<br>0.277 | (+)<br><0.001 |
|  | 24 | Oral and oromucosal-fair/good | 50 | 0.86 |  |  |  |  |  |  | (+)<br>0.230 | (+)<br><0.001 |
|  | 25 | Oral-fair/good | 35 | 0.87 |  |  |  |  |  |  | (+)<br>0.005 | (+)<br><0.001 |

\*Reference group for route of administration was “oral”, for CBD formulation was “Epidiolex”, and for diet was “fast” status.
†The number of female participants was divided by the total participants, and the result ratio was a number between 0 and 1, which was included in the model.
This table does not include regression coefficients since they were in log scale and not interpretable in terms of effect size. Hereby only the sig of regression coefficients, i.e. negative or positive, and statistical significance level are reported.

## Discussion

In this review, we aimed to provide an updated systematic assessment of available evidence on the PK of CBD, provide comparable PK parameters from different studies on the same scale, and explore the impact of different relevant factors on PK outcomes. There was considerable heterogeneity in the available PK data for CBD both in terms of the study conduct and reported outcomes. Nevertheless, despite the heterogeneity and quality aspects, several meaningful patterns emerged for factors expected to influence the pharmacokinetic outcomes of CBD including the route of administration, dose, formulation, diet status, sex ratio, and study duration.

For the parameters related to bioavailability, i.e. Cmax, AUC0-t, AUC0-inf, it appeared that inhalation and oral administration had comparable outcomes. However, oromucosal administration consistently resulted in a lower bioavailability than oral administration, which is in line with a previous systematic review ^3^ and some of the within-study comparisons using a similar dose for both routes of administration ^31,35^. There are though other within-study comparisons indicating that bioavailability was comparable between oral and oromucosal administration ^16^, or that oral administration resulted in lower bioavailability ^49^. Different CBD formulations for oral and oromucosal administration could potentially explain these within-study inconsistencies, at least in part. However, due to the low CBD doses used particularly in oromucosal trial arms, comparability is limited and reliable interpretation warrants additional studies. Regarding rate of absorption, as would be expected, inhalation resulted in the lower Tmax/faster absorption compared to oral administration. There was a lack of significant difference between oromucosal and oral administration regarding absorption rate, which is in line with a previous systematic review ^3^ as well as some of the direct within-study comparisons _16,31,35_.

Given the general low bioavailability of oral administration of cannabinoids, recent years have seen an increase in CBD nanoformulations in hope of increasing absorption and bioavailability through the oral and oromucosal administration routes. In the regression models for Tmax, model fit was noticeably improved by accounting for formulation. Nanotech and oil-based formulations were associated with lower Tmax than the Epidiolex formulation indicating that they would have a faster onset of action. To our knowledge, there has been no published direct side-by-side comparison of Epidiolex with nanotech or oil-based formulations. In this review, due to the methodological considerations that are discussed in the statistics section and supplementary methods, only certain formulations could be included in the regression models for Tmax. Therefore, we could not explore other formulations or other PK parameters, thus limiting interpretation of different formulations on their comparable bioavailability for CBD.

As expected, higher CBD dose was consistently associated with higher bioavailability in all the models for Cmax, AUC0-t, and AUC0-inf. The information provided in Table 3 and Figure 2 serve as a useful reference to help predict CBD dose to reach a certain serum level and potential clinical effect for specific conditions. While various CBD formulations have been used in lower doses, all the studies with a CBD dose of >100mg were only conducted with Epidiolex. As such, there is still a lack of PK information about CBD doses expected to be more aligned with a ‘medicinal’ range.

Food consumption can influence bioavailability of many medications, especially lipophilic compounds like CBD through increased transit time and lymphatic absorption in the intestines. We observed that fed condition was associated with a higher bioavailability of CBD, but had no significant impact in the time to reach maximum serum concentrations, compared to the fasted status. Specifically, we observed a higher Cmax and AUC0-t across all the models for fed condition compared to fast status, which is in line with within-study direct comparisons ^12,19,34,40^. The lack of a significant effect of diet status on Tmax, is in line with two multiple-arm comprehensive studies ^19,34^, but inconsistent with two other studies where Tmax was considerably longer in the fed group ^12,40^. Although we would expect a significant effect of diet on AUC0-inf, both theoretically and based on within-study comparisons ^19,34,40^, we did not detect such an effect in our models. This could be in part attributed to the lower number of studies that reported AUC0-inf and thus lower power of these models.

The importance of sex for drug bioavailability and clearance could also have significant implications, including a probably lower required dose of CBD in females to reach a certain blood level and clinical effect. There are noted sex differences in CYP450 family of enzymes which contribute to the metabolism of CBD ^51^. Also the literature suggests slower clearance in females compared to males ^52,53^. We found that for the PK studies in which sex ratio was reported, a higher female ratio was associated with a faster absorption and higher maximum concentrations through oral administration, evident with lower Tmax and higher Cmax respectively. These findings are consistent with direct within-study CBD PK comparisons showing higher Cmax and lower Tmax in female participants compared with males ^26,45^. A recent PK study with very low doses of CBD reported no effect for sex on Tmax, but female sex was associated with significantly higher Cmax and AUC0-t ^14^. A higher Cmax and lower T_1/2_ with oral administration would mathematically be expected to be associated with a higher AUC0-t and AUC0-inf and also in line with direct within-study sex comparisons ^14,26,45^.

However, this relationship was not evident in our analysis perhaps due to the few studies conducted with females. Well-powered studies with clinically relevant higher doses in males and females are still needed.

As expected, duration of PK session was a determinant of overall bioavailability and clearance of CBD with implications for designing future studies. A longer duration of PK session was associated with a higher AUC0-inf in all models, and a higher AUC0-t in models that included data from all routes of administration of CBD. A higher duration of the PK session was also consistently associated with a longer T_1/2_ across all models. In line with this later finding, the literature suggests that cannabinoids in humans may need extended durations to adequately determine cannabinoid half-lives, given that the terminal half-life of cannabinoids is longer than the initial half-life ^6^. Indeed, half-lives of longer than 60h were reported for Epidiolex with a PK duration of 168h ^23^.

Certain study characteristics could limit cross-study comparisons, including different reporting scales, considerable heterogeneity of study conditions, and lack of clarity in details of some formulations, particularly for commercialized products. Presence of other cannabinoids in the preparations is also an important consideration. For example, THC could hypothetically influence CBD PK at the level of shared metabolic pathways, although, to the best of authors’ knowledge, there are no human PK study to date that has systematically compared CBD bioavailability with and without THC. In addition, CBDA in some of the preparations could readily convert to CBD in the human body and alter pharmacokinetic outcomes. There were also certain limitations related to the methods of this review. Comprehensive quality assessment of PK studies requires a specifically designed assessment tool beyond the ones used for general before-after studies. Although there have been efforts to introduce such quality assessment tools ^54^, there is still no commonly accepted one available. In addition to the items covered by the NIHLB tool, pre-registration of study protocol as a clinical trial, the potential effect of other constituents of the administered product on the PK outcomes, sufficient length of study, accounting for the potential effect of important covariates like sex and body mass, would be important considerations. In regard to the statistical methods, the log-transformed regression coefficients could not be interpreted in terms of effect size, limiting our prediction of each factor’s impact on PK parameters. Finally, we did not include the PK outcomes of CBD metabolites reported by some of the included studies since it metabolism is affected by multiple factors thus requiring a focused and extensive exploration that was outside the scope of this review. Understanding the pharmacokinetics of CBD metabolites can provide further insights into the CBD half/life and sex differences.

## Conclusion

We provided an updated overview of the current status of evidence of pharmacokinetics of CBD. In exploring how different factors potentially influenced the PK outcomes of CBD, consuming food while taking CBD, female sex, and oral administration were associated with higher bioavailability. Recommendations for future research would mainly concern conducting original systematic studies to elucidate the impact of biological sex and different formulations in single studies with multiple arms. It would also be beneficial for future studies to examine clinically relevant doses of CBD, e.g., >200mg, given the increasing number of such preparations on the market and their potential application in clinical practice. Finally, reporting PK parameters in both arithmetic and geometric scales would help improve comparisons across studies and would also enhance knowledge aggregation and replicability.

## Data Availability

All data produced in the present study are available upon reasonable request to the authors

## Acknowledgements

None

## Supplementary Material

Supplementary methods, sections A-C

Supplementary Table 1

Supplementary Figure 1

---

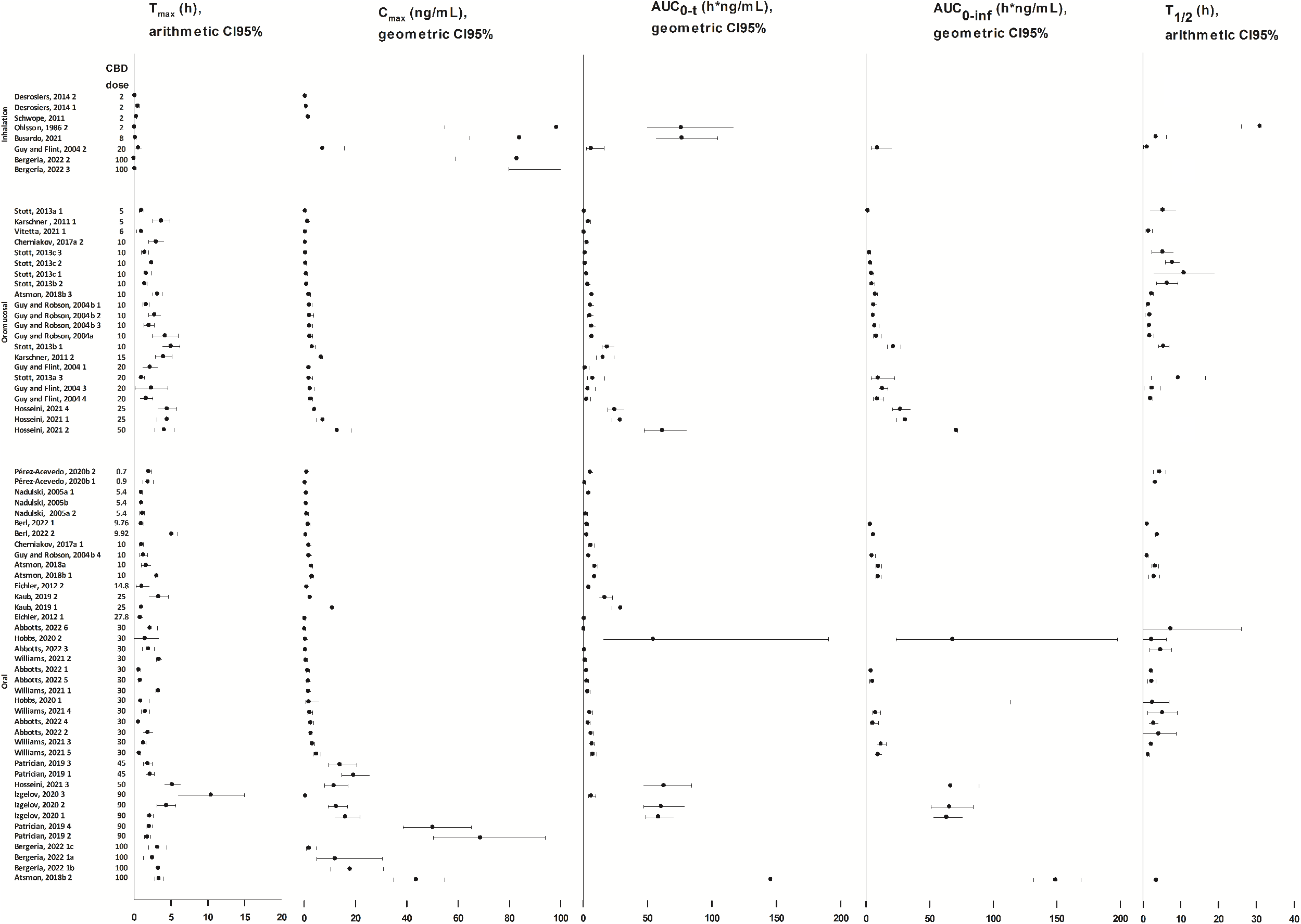

**Supplementary Table 1.**
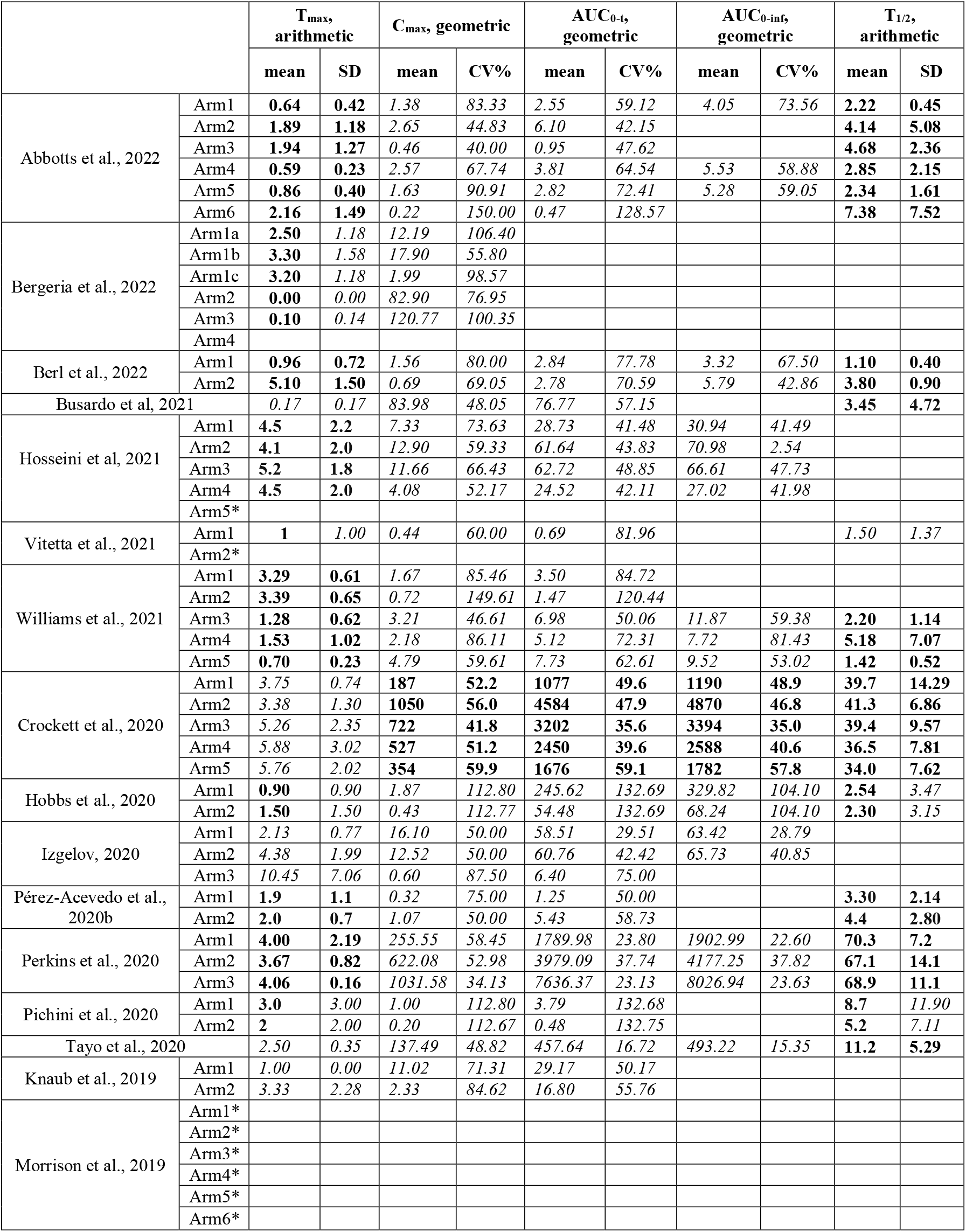
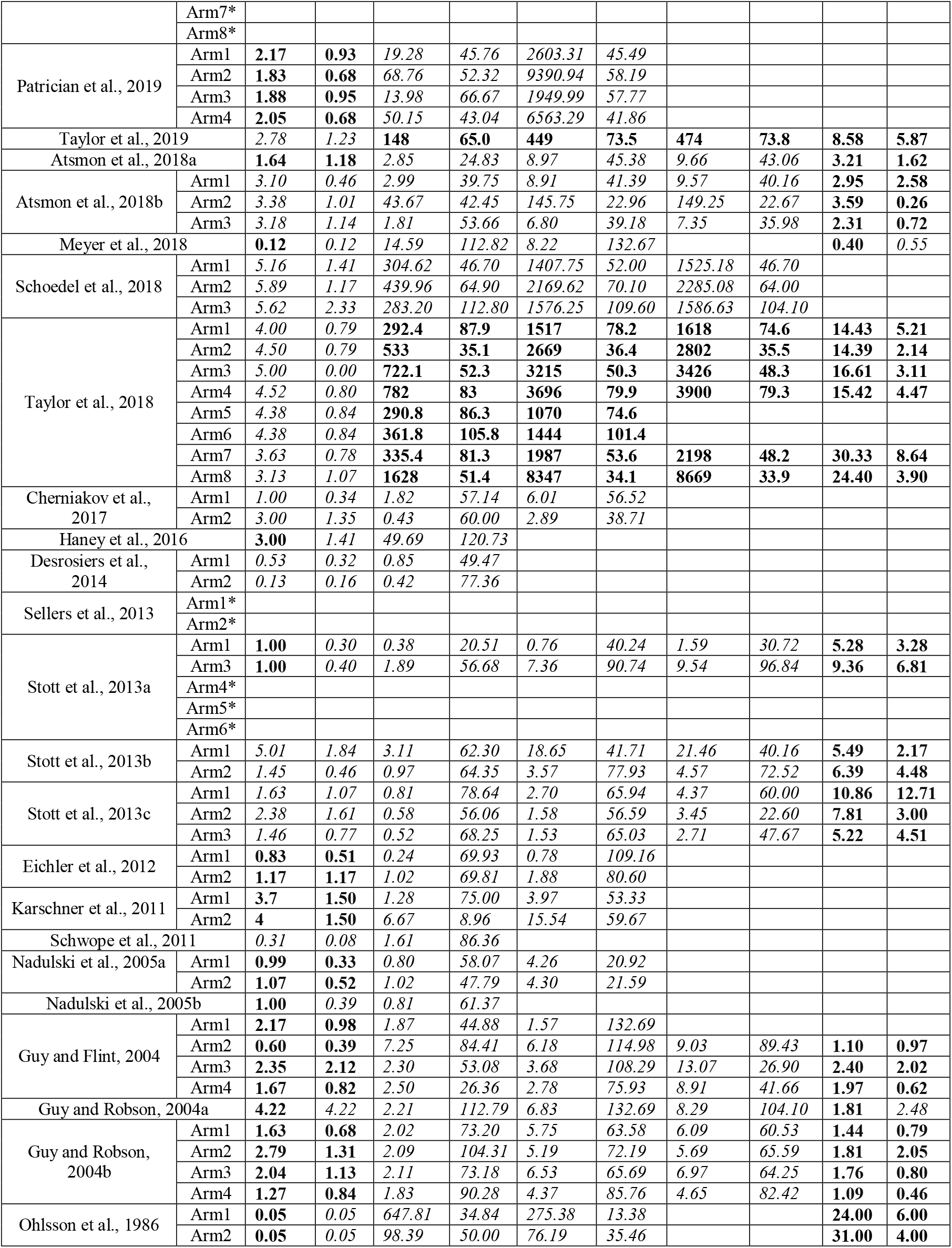
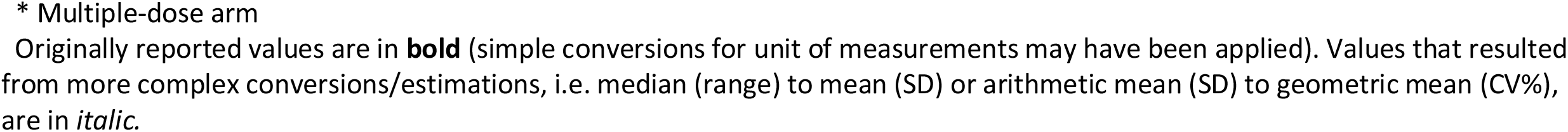
Pharmacokinetic parameters of cannabidiol studies in accordance with their order in Table 1

|  |  | T<sub>max</sub>,<br>arithmetic |  | C<sub>max</sub>, geometric |  | AUC<sub>0-t<sub>s</sub></sub>,<br>geometric |  | AUC<sub>0-inf</sub>,<br>geometric |  | T<sub>1/2</sub>,<br>arithmetic |  |
| --- | --- | --- | --- | --- | --- | --- | --- | --- | --- | --- | --- |
|  |  | mean | SD | mean | CV% | mean | CV% | mean | CV% | mean | SD |
| Abbotts et al., 2022 | Arm1 | 0.64 | 0.42 | 1.38 | 83.33 | 2.55 | 59.12 | 4.05 | 73.56 | 2.22 | 0.45 |
|  | Arm2 | 1.89 | 1.18 | 2.65 | 44.83 | 6.10 | 42.15 |  |  | 4.14 | 5.08 |
|  | Arm3 | 1.94 | 1.27 | 0.46 | 40.00 | 0.95 | 47.62 |  |  | 4.68 | 2.36 |
|  | Arm4 | 0.59 | 0.23 | 2.57 | 67.74 | 3.81 | 64.54 | 5.53 | 58.88 | 2.85 | 2.15 |
|  | Arm5 | 0.86 | 0.40 | 1.63 | 90.91 | 2.82 | 72.41 | 5.28 | 59.05 | 2.34 | 1.61 |
|  | Arm6 | 2.16 | 1.49 | 0.22 | 150.00 | 0.47 | 128.57 |  |  | 7.38 | 7.52 |
| Bergeria et al., 2022 | Arm1a | 2.50 | 1.18 | 12.19 | 106.40 |  |  |  |  |  |  |
|  | Arm1b | 3.30 | 1.58 | 17.90 | 55.80 |  |  |  |  |  |  |
|  | Arm1c | 3.20 | 1.18 | 1.99 | 98.57 |  |  |  |  |  |  |
|  | Arm2 | 0.00 | 0.00 | 82.90 | 76.95 |  |  |  |  |  |  |
|  | Arm3 | 0.10 | 0.14 | 120.77 | 100.35 |  |  |  |  |  |  |
|  | Arm4 |  |  |  |  |  |  |  |  |  |  |
| Berl et al., 2022 | Arm1 | 0.96 | 0.72 | 1.56 | 80.00 | 2.84 | 77.78 | 3.32 | 67.50 | 1.10 | 0.40 |
|  | Arm2 | 5.10 | 1.50 | 0.69 | 69.05 | 2.78 | 70.59 | 5.79 | 42.86 | 3.80 | 0.90 |
| Busardo et al, 2021 |  | 0.17 | 0.17 | 83.98 | 48.05 | 76.77 | 57.15 |  |  | 3.45 | 4.72 |
| Hosseini et al, 2021 | Arm1 | 4.5 | 2.2 | 7.33 | 73.63 | 28.73 | 41.48 | 30.94 | 41.49 |  |  |
|  | Arm2 | 4.1 | 2.0 | 12.90 | 59.33 | 61.64 | 43.83 | 70.98 | 2.54 |  |  |
|  | Arm3 | 5.2 | 1.8 | 11.66 | 66.43 | 62.72 | 48.85 | 66.61 | 47.73 |  |  |
|  | Arm4 | 4.5 | 2.0 | 4.08 | 52.17 | 24.52 | 42.11 | 27.02 | 41.98 |  |  |
|  | Arm5* |  |  |  |  |  |  |  |  |  |  |
| Vitetta et al., 2021 | Arm1 | 1 | 1.00 | 0.44 | 60.00 | 0.69 | 81.96 |  |  | 1.50 | 1.37 |
|  | Arm2* |  |  |  |  |  |  |  |  |  |  |
| Williams et al., 2021 | Arm1 | 3.29 | 0.61 | 1.67 | 85.46 | 3.50 | 84.72 |  |  |  |  |
|  | Arm2 | 3.39 | 0.65 | 0.72 | 149.61 | 1.47 | 120.44 |  |  |  |  |
|  | Arm3 | 1.28 | 0.62 | 3.21 | 46.61 | 6.98 | 50.06 | 11.87 | 59.38 | 2.20 | 1.14 |
|  | Arm4 | 1.53 | 1.02 | 2.18 | 86.11 | 5.12 | 72.31 | 7.72 | 81.43 | 5.18 | 7.07 |
|  | Arm5 | 0.70 | 0.23 | 4.79 | 59.61 | 7.73 | 62.61 | 9.52 | 53.02 | 1.42 | 0.52 |
| Crockett et al., 2020 | Arm1 | 3.75 | 0.74 | 187 | 52.2 | 1077 | 49.6 | 1190 | 48.9 | 39.7 | 14.29 |
|  | Arm2 | 3.38 | 1.30 | 1050 | 56.0 | 4584 | 47.9 | 4870 | 46.8 | 41.3 | 6.86 |
|  | Arm3 | 5.26 | 2.35 | 722 | 41.8 | 3202 | 35.6 | 3394 | 35.0 | 39.4 | 9.57 |
|  | Arm4 | 5.88 | 3.02 | 527 | 51.2 | 2450 | 39.6 | 2588 | 40.6 | 36.5 | 7.81 |
|  | Arm5 | 5.76 | 2.02 | 354 | 59.9 | 1676 | 59.1 | 1782 | 57.8 | 34.0 | 7.62 |
| Hobbs et al., 2020 | Arm1 | 0.90 | 0.90 | 1.87 | 112.80 | 245.62 | 132.69 | 329.82 | 104.10 | 2.54 | 3.47 |
|  | Arm2 | 1.50 | 1.50 | 0.43 | 112.77 | 54.48 | 132.69 | 68.24 | 104.10 | 2.30 | 3.15 |
| Izgelov, 2020 | Arm1 | 2.13 | 0.77 | 16.10 | 50.00 | 58.51 | 29.51 | 63.42 | 28.79 |  |  |
|  | Arm2 | 4.38 | 1.99 | 12.52 | 50.00 | 60.76 | 42.42 | 65.73 | 40.85 |  |  |
|  | Arm3 | 10.45 | 7.06 | 0.60 | 87.50 | 6.40 | 75.00 |  |  |  |  |
| Pérez-Acevedo et al., 2020b | Arm1 | 1.9 | 1.1 | 0.32 | 75.00 | 1.25 | 50.00 |  |  | 3.30 | 2.14 |
|  | Arm2 | 2.0 | 0.7 | 1.07 | 50.00 | 5.43 | 58.73 |  |  | 4.4 | 2.80 |
| Perkins et al., 2020 | Arm1 | 4.00 | 2.19 | 255.55 | 58.45 | 1789.98 | 23.80 | 1902.99 | 22.60 | 70.3 | 7.2 |
|  | Arm2 | 3.67 | 0.82 | 622.08 | 52.98 | 3979.09 | 37.74 | 4177.25 | 37.82 | 67.1 | 14.1 |
|  | Arm3 | 4.06 | 0.16 | 1031.58 | 34.13 | 7636.37 | 23.13 | 8026.94 | 23.63 | 68.9 | 11.1 |
| Pichini et al., 2020 | Arm1 | 3.0 | 3.00 | 1.00 | 112.80 | 3.79 | 132.68 |  |  | 8.7 | 11.90 |
|  | Arm2 | 2 | 2.00 | 0.20 | 112.67 | 0.48 | 132.75 |  |  | 5.2 | 7.11 |
| Tayo et al., 2020 |  | 2.50 | 0.35 | 137.49 | 48.82 | 457.64 | 16.72 | 493.22 | 15.35 | 11.2 | 5.29 |
| Knaub et al., 2019 | Arm1 | 1.00 | 0.00 | 11.02 | 71.31 | 29.17 | 50.17 |  |  |  |  |
|  | Arm2 | 3.33 | 2.28 | 2.33 | 84.62 | 16.80 | 55.76 |  |  |  |  |
| Morrison et al., 2019 | Arm1* |  |  |  |  |  |  |  |  |  |  |
|  | Arm2* |  |  |  |  |  |  |  |  |  |  |
|  | Arm3* |  |  |  |  |  |  |  |  |  |  |
|  | Arm4* |  |  |  |  |  |  |  |  |  |  |
|  | Arm5* |  |  |  |  |  |  |  |  |  |  |
|  | Arm6* |  |  |  |  |  |  |  |  |  |  |
|  | Arm7* |  |  |  |  |  |  |  |  |  |  |
|  | Arm8* |  |  |  |  |  |  |  |  |  |  |
| Patrician et al., 2019 | Arm1 | **2.17** | **0.93** | 19.28 | 45.76 | 2603.31 | 45.49 |  |  |  |  |
|  | Arm2 | **1.83** | **0.68** | 68.76 | 52.32 | 9390.94 | 58.19 |  |  |  |  |
|  | Arm3 | **1.88** | **0.95** | 13.98 | 66.67 | 1949.99 | 57.77 |  |  |  |  |
|  | Arm4 | **2.05** | **0.68** | 50.15 | 43.04 | 6563.29 | 41.86 |  |  |  |  |
| Taylor et al., 2019 |  | 2.78 | 1.23 | **148** | **65.0** | **449** | **73.5** | **474** | **73.8** | **8.58** | **5.87** |
| Atsmon et al., 2018a |  | **1.64** | **1.18** | 2.85 | 24.83 | 8.97 | 45.38 | 9.66 | 43.06 | **3.21** | **1.62** |
| Atsmon et al., 2018b | Arm1 | 3.10 | 0.46 | 2.99 | 39.75 | 8.91 | 41.39 | 9.57 | 40.16 | **2.95** | **2.58** |
|  | Arm2 | 3.38 | 1.01 | 43.67 | 42.45 | 145.75 | 22.96 | 149.25 | 22.67 | **3.59** | **0.26** |
|  | Arm3 | 3.18 | 1.14 | 1.81 | 53.66 | 6.80 | 39.18 | 7.35 | 35.98 | **2.31** | **0.72** |
| Meyer et al., 2018 |  | **0.12** | 0.12 | 14.59 | 112.82 | 8.22 | 132.67 |  |  | **0.40** | 0.55 |
| Schoedel et al., 2018 | Arm1 | 5.16 | 1.41 | 304.62 | 46.70 | 1407.75 | 52.00 | 1525.18 | 46.70 |  |  |
|  | Arm2 | 5.89 | 1.17 | 439.96 | 64.90 | 2169.62 | 70.10 | 2285.08 | 64.00 |  |  |
|  | Arm3 | 5.62 | 2.33 | 283.20 | 112.80 | 1576.25 | 109.60 | 1586.63 | 104.10 |  |  |
| Taylor et al., 2018 | Arm1 | 4.00 | 0.79 | **292.4** | **87.9** | **1517** | **78.2** | **1618** | **74.6** | **14.43** | **5.21** |
|  | Arm2 | 4.50 | 0.79 | **533** | **35.1** | **2669** | **36.4** | **2802** | **35.5** | **14.39** | **2.14** |
|  | Arm3 | 5.00 | 0.00 | **722.1** | **52.3** | **3215** | **50.3** | **3426** | **48.3** | **16.61** | **3.11** |
|  | Arm4 | 4.52 | 0.80 | **782** | **83** | **3696** | **79.9** | **3900** | **79.3** | **15.42** | **4.47** |
|  | Arm5 | 4.38 | 0.84 | **290.8** | **86.3** | **1070** | **74.6** |  |  |  |  |
|  | Arm6 | 4.38 | 0.84 | **361.8** | **105.8** | **1444** | **101.4** |  |  |  |  |
|  | Arm7 | 3.63 | 0.78 | **335.4** | **81.3** | **1987** | **53.6** | **2198** | **48.2** | **30.33** | **8.64** |
|  | Arm8 | 3.13 | 1.07 | **1628** | **51.4** | **8347** | **34.1** | **8669** | **33.9** | **24.40** | **3.90** |
| Cherniakov et al., 2017 | Arm1 | 1.00 | 0.34 | 1.82 | 57.14 | 6.01 | 56.52 |  |  |  |  |
|  | Arm2 | 3.00 | 1.35 | 0.43 | 60.00 | 2.89 | 38.71 |  |  |  |  |
| Haney et al., 2016 |  | **3.00** | 1.41 | 49.69 | 120.73 |  |  |  |  |  |  |
| Desrosiers et al., 2014 | Arm1 | 0.53 | 0.32 | 0.85 | 49.47 |  |  |  |  |  |  |
|  | Arm2 | 0.13 | 0.16 | 0.42 | 77.36 |  |  |  |  |  |  |
| Sellers et al., 2013 | Arm1* |  |  |  |  |  |  |  |  |  |  |
|  | Arm2* |  |  |  |  |  |  |  |  |  |  |
| Stott et al., 2013a | Arm1 | **1.00** | 0.30 | 0.38 | 20.51 | 0.76 | 40.24 | 1.59 | 30.72 | **5.28** | **3.28** |
|  | Arm3 | **1.00** | 0.40 | 1.89 | 56.68 | 7.36 | 90.74 | 9.54 | 96.84 | **9.36** | **6.81** |
|  | Arm4* |  |  |  |  |  |  |  |  |  |  |
|  | Arm5* |  |  |  |  |  |  |  |  |  |  |
|  | Arm6* |  |  |  |  |  |  |  |  |  |  |
| Stott et al., 2013b | Arm1 | 5.01 | 1.84 | 3.11 | 62.30 | 18.65 | 41.71 | 21.46 | 40.16 | **5.49** | **2.17** |
|  | Arm2 | 1.45 | 0.46 | 0.97 | 64.35 | 3.57 | 77.93 | 4.57 | 72.52 | **6.39** | **4.48** |
| Stott et al., 2013c | Arm1 | 1.63 | 1.07 | 0.81 | 78.64 | 2.70 | 65.94 | 4.37 | 60.00 | **10.86** | **12.71** |
|  | Arm2 | 2.38 | 1.61 | 0.58 | 56.06 | 1.58 | 56.59 | 3.45 | 22.60 | **7.81** | **3.00** |
|  | Arm3 | 1.46 | 0.77 | 0.52 | 68.25 | 1.53 | 65.03 | 2.71 | 47.67 | **5.22** | **4.51** |
| Eichler et al., 2012 | Arm1 | **0.83** | **0.51** | 0.24 | 69.93 | 0.78 | 109.16 |  |  |  |  |
|  | Arm2 | **1.17** | **1.17** | 1.02 | 69.81 | 1.88 | 80.60 |  |  |  |  |
| Karschner et al., 2011 | Arm1 | **3.7** | **1.50** | 1.28 | 75.00 | 3.97 | 53.33 |  |  |  |  |
|  | Arm2 | **4** | **1.50** | 6.67 | 8.96 | 15.54 | 59.67 |  |  |  |  |
| Schwope et al., 2011 |  | 0.31 | 0.08 | 1.61 | 86.36 |  |  |  |  |  |  |
| Nadulski et al., 2005a | Arm1 | **0.99** | **0.33** | 0.80 | 58.07 | 4.26 | 20.92 |  |  |  |  |
|  | Arm2 | **1.07** | **0.52** | 1.02 | 47.79 | 4.30 | 21.59 |  |  |  |  |
| Nadulski et al., 2005b |  | **1.00** | 0.39 | 0.81 | 61.37 |  |  |  |  |  |  |
| Guy and Flint, 2004 | Arm1 | **2.17** | **0.98** | 1.87 | 44.88 | 1.57 | 132.69 |  |  |  |  |
|  | Arm2 | **0.60** | **0.39** | 7.25 | 84.41 | 6.18 | 114.98 | 9.03 | 89.43 | **1.10** | **0.97** |
|  | Arm3 | **2.35** | **2.12** | 2.30 | 53.08 | 3.68 | 108.29 | 13.07 | 26.90 | **2.40** | **2.02** |
|  | Arm4 | **1.67** | **0.82** | 2.50 | 26.36 | 2.78 | 75.93 | 8.91 | 41.66 | **1.97** | **0.62** |
| Guy and Robson, 2004a |  | **4.22** | 4.22 | 2.21 | 112.79 | 6.83 | 132.69 | 8.29 | 104.10 | **1.81** | 2.48 |
| Guy and Robson, 2004b | Arm1 | **1.63** | **0.68** | 2.02 | 73.20 | 5.75 | 63.58 | 6.09 | 60.53 | **1.44** | **0.79** |
|  | Arm2 | **2.79** | **1.31** | 2.09 | 104.31 | 5.19 | 72.19 | 5.69 | 65.59 | **1.81** | **2.05** |
|  | Arm3 | **2.04** | **1.13** | 2.11 | 73.18 | 6.53 | 65.69 | 6.97 | 64.25 | **1.76** | **0.80** |
|  | Arm4 | **1.27** | **0.84** | 1.83 | 90.28 | 4.37 | 85.76 | 4.65 | 82.42 | **1.09** | **0.46** |
| Ohlsson et al., 1986 | Arm1 | **0.05** | 0.05 | 647.81 | 34.84 | 275.38 | 13.38 |  |  | **24.00** | **6.00** |
|  | Arm2 | **0.05** | 0.05 | 98.39 | 50.00 | 76.19 | 35.46 |  |  | **31.00** | **4.00** |

\* Multiple-dose arm
Originally reported values are in **bold** (simple conversions for unit of measurements may have been applied). Values that resulted from more complex conversions/estimations, i.e. median (range) to mean (SD) or arithmetic mean (SD) to geometric mean (CV%), are in *italic*.

## Pharmacokinetics of Cannabidiol

### A systematic meta-regression analysis to guide clinical trials

#### Section A_ Quality Assessment

Authors of this review chose a more precise approach to the overall rating of NHLBI tool toward a more objective unified replicable assessment. Given the unique characteristics of PK studies and their outcomes, it was decided that certain items from the tool bear more weight in determining the overall risk of bias and quality of the study; question 2 is concerned with description of study population, question 5 with sample size, and question 6 with description of the intervention. This difference in weighting was applied through the following logic:

-A study is rated as Poor if the answer to more than four questions is either NR/CD/NO, or if the answer to exactly four questions is NR/CD/NO and one of them is among questions number 2, 5, or 6.

-A study is rated as Fair, if the answer to three or four questions other than 2,5, or 6 is NR/CD/NO, or if the answer to two questions is NR/CD/NO and one of them is among questions number 2, 5, or 6.

-A study is rated as good if the answer to two or less than two questions is NR/CD/NO and none of them is among questions number 2, 5, or 6.

#### Section B_ Estimation/conversion of PK values

The formulas that are discussed in this section are based on highly cited published work and widely accepted methods by expert statisticians (Wan et al., 2014; Higgins et al., 2008) and in accordance with available guidelines (Cochrane Handbook 6.3, 2022). Meanwhile, they have been rarely used in reviews of PK studies which could be in part due to the complexity and time-consuming nature of such an approach. Hereby, we try to provide a simplified guide for recruiting these methods. We also showcase the level of accuracy of the results after arithmetic to geometric conversion, using arithmetic outcomes from Morrison et al., 2019, where authors reported both arithmetic and geometric values for 3 medication arms calculated originally from individual-participant data. More comprehensive showcases and evaulation of these methods are available elsewhere (Wan et al., 2014; Higgins et al., 2008).

#### Methods

For PK values reported as median (range), formula number 3 and 7 from Wan et al., 2014 were used to estimate arithmetic mean and SD respectively as following:

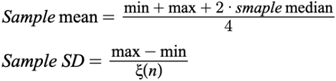

where *ξ*(*n*) comes from Table1 in Wan et al., 2014, and it’s value depends on the sample size.

For PK values reported as arithmetic mean (SD), a series of formulas presented on pages 6073-6075 from Higgins et al., 2008 was used, which are revised for simplicity and understandability (without any change to the original formula):

Step1→ a z^−^ and S_z_ value are estimated as mean and standard deviation of log-transformed measurements respectively, using the following formula:

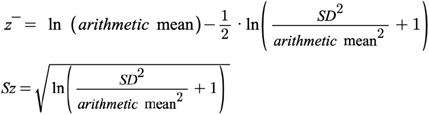

Step2→ geometric mean can be estimated from arithmetic mean using the following formula:

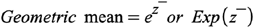

Step3→ calculation of geometric CV% from arithmetic data is not directly discussed in Higgins et al., 2008, and the authors of this review could not find an original paper directly discussing this estimation. Meanwhile, several statistical experts has further discussed, applied, and modified the concepts of “geometric measures of dispersion” proposed by Kirkwood et al., 1979. The final outcome has been a currently commonly used form (e.g. in SAS software) as geometric coefficient of variation= sqrt(exp((standard deviation of log-transformed data) ^2^)-1) (https://blogs.sas.com/content/iml/2019/10/02/geometric-mean-deviation-cv-sas.html) where “standard deviation of log-transformed data” could be replaced by Higgins et al definition of Sz. As such, the following formula can be used for estimation of geometric CV%:

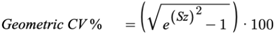

Step4→ for the purpose of meta-regression analysis, z^−^ (CI95%) was calculated for all the values of all PK parameters in single dose arms, and was used as input for linear meta-regression analysis, using Higgins et al 2008 methods. It is noteworthy that z^−^ (CI95%) is in fact log-normal-transformed version of geometric mean (geometric confidence interval). Two possible scenarios are as following:

Scenario1-if data is reported as arithmetic mean (SD), then

*Log-transformed geometric mean*= *z^−^*

*Log-transformed geometric CI95%* = 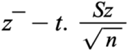 *to* 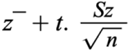

*t-score can be calculated using any statistical software or any of the numerous online t-score calculators

Scenario 2-if data is already reported as geometric mean (CV%), then

*Log-transformed geometric mean*= *z^−^= ln (Geometric mean)*

*Log-transformed geometric CI95%* = 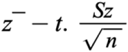 *to* 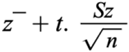

*t-score can be calculated using any statistical software or any of the numerous online t-score calculators

*Sz in here cannot be calculated using the formula discussed before, since that was based on arithmetic data. Given the previous discussion of the work by Kirkwood et al 1997, and currently accepted methods, the following formula can be used to estimate Sz from geometric CV%:

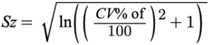

Step5→ for demonstration purposes, like Suppl Fig1, geometric CI95% can be simply estimated as following:

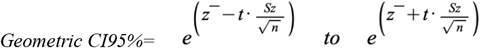

#### Showcase

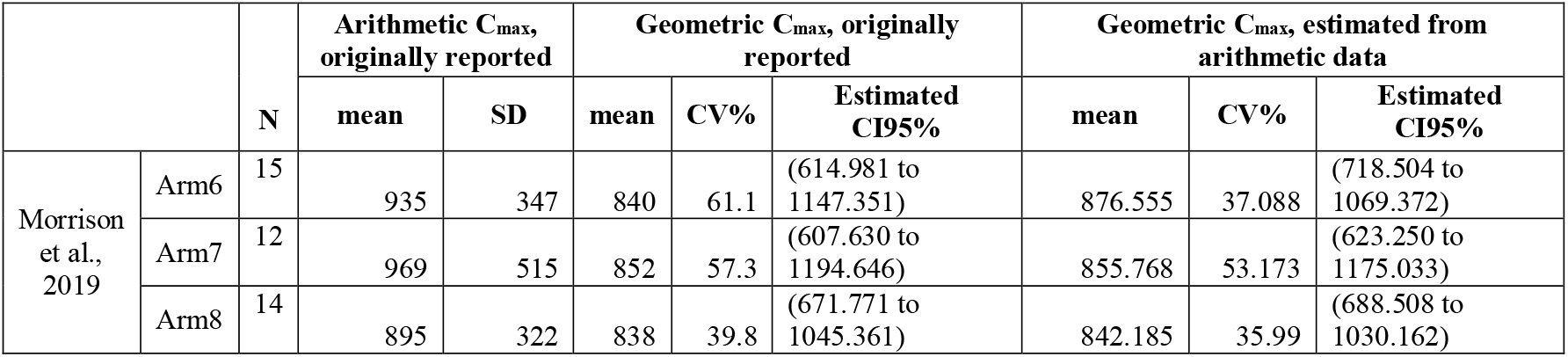

|  |  | N | Arithmetic AUC<sub>0-t</sub>, originally reported |  | Geometric AUC<sub>0-t</sub>, originally reported |  |  | Geometric AUC<sub>0-t</sub>, estimated from arithmetic data |  |  |
| --- | --- | --- | --- | --- | --- | --- | --- | --- | --- | --- |
|  |  |  | mean | SD | mean | CV% | Estimated CI95% | mean | CV% | Estimated CI95% |
| Morrison et al., 2019 | Arm6 | 15 | 3720 | 1060 | 3500 | 42.6 | (2792.110 to 4387.362) | 3575.999 | 28.451 | (3064.009 to 4173.542) |
|  | Arm7 | 12 | 3880 | 1260 | 3690 | 34.2 | (2988.233 to 4556.572) | 3688.591 | 32.513 | (3015.697 to 4511.628) |
|  | Arm8 | 14 | 3740 | 1110 | 3560 | 35.8 | (2913.755 to 4349.575) | 3586.743 | 29.727 | (3032.098 to 4242.846) |

## Section C_ Considerations for meta-regression modeling

For each PK parameter, only the variables that were theoretically relevant based on the pharmacokinetics literature were included in the model, e.g. only duration of the PK session and female ratio but not CBD dose or route of administration were included in the model for T1/2. In order to prevent overfitting, high collinearity, or exclusively correlated variables that could result in misinterpretation, the following considerations were applied: - if coefficient of correlation was more than 0.6 for any of the two independent variables, only the one with more robust available evidence was included in the model. -any independent categorical variable that was included had to have all the different possible values present across other categorical variables, to prevent exclusive correlations. e.g. for “abstinence status” to be included in the model beside the “route of administration”, there had to be different medication arms with both abstinent participants and non-abstinent participants in all the routes of administration that were included in the model, e.g. oromucosal {abstinent, non-abstinent}, oral {abstinent, non-abstinent}. In this particular example, since there were only one non-abstinent medication arm among oral medication administration arms and none among oromucosal administration arms, this variable was not included in any of the models. Also, since all the medication arms with CBD dose of >100mg were conducted with Epidiolex, and some formulations were exclusive to either oral or oromucosal adminsitartion, we avoided including both the CBD dose and formulation simultaneously in the models for Cmax and AUC and therefore only included CBD dose. However, we could include the CBD formulation in the model for Tmax for oral administration arms, given that CBD dose was not conceptually relevant to Tmax to be included in that model, and thus there was no concern about dose-formulation collinearity for Tmax.

